# Psilocybin treatment for symptoms of depression: a living systematic review, meta-analysis, and data resource

**DOI:** 10.1101/2025.08.13.25333530

**Authors:** S. Parker Singleton, Brooke L. Sevchik, Analiese Lahey, Pim Cuijpers, Mathias Harrer, Megan T. Jones, Sandeep M. Nayak, Eric C. Strain, Simon N. Vandekar, Robert H. Dworkin, J. Cobb Scott, Theodore D. Satterthwaite

## Abstract

**Importance:** Depression is a major cause of disability worldwide, motivating substantial interest in psilocybin as a potential treatment.

**Objective:** To conduct a systematic review and meta-analysis of psilocybin’s impact on depressive symptoms and provide a living open data resource.

**Data Sources:** PubMed, Embase, Scopus, Web of Science, and PsycINFO retrieved by a systematic search up to July 1, 2025.

**Study Selection:** We included randomized controlled trials of psilocybin or psilocybin-assisted therapy compared against a placebo or waitlist condition.

**Data Extraction and Synthesis:** Data extraction was completed independently by two extractors. A random-effects meta-analysis was used to synthesize data. Risk of bias was assessed with Cochrane’s RoB 2.0 tool.

**Main Outcomes and Measures:** The main outcome was the standardized mean difference (Hedges’ *g*) in depression scores at the primary study endpoint.

**Results:** Twelve studies comprising 711 participants were included in the database, with nine of those studies (n = 529) included in our primary model. Of the nine studies included in the primary model, two had a high risk of bias, four had some concerns, while three had a low risk of bias. Compared to control conditions, psilocybin showed a greater reduction in depression scores, with a pooled Hedges’ *g* = –0.91 (95% CI, [-1.35; –0.48]; *k* = 9; *p* = 0.0013, *I*^2^ = 58.1%, *tau*^2^ *=* 0.13, *n* = 501). Sensitivity analyses revealed robust effects consistent with the primary model across a variety of design parameters and analysis choices, while also suggesting that waitlist control and crossover design studies contribute a large amount of heterogeneity to the primary model. Meta-regression revealed that psilocybin’s effects were rapid and consistent over several weeks (intercept = –0.92 [-1.26; –0.58], *p* < 0.0001; slope = 0.0009 [-0.0023; 0.0041], *p* = 0.57).

**Conclusions and Relevance:** This systematic review and meta-analysis suggests that psilocybin-assisted therapy results in substantial decreases in depressive symptoms across studies to date. However, many studies have small sample sizes or risk of bias. This living systematic review, meta-analysis, database, and online dashboard will continue to be updated as evidence emerges, providing a valuable resource for researchers in a rapidly evolving field.

**Key Points:** *Question:* What is the efficacy of psilocybin or psilocybin-assisted therapy for depressive symptoms?

*Findings:* In this living systematic review and meta-analysis, the initial evidence suggests that psilocybin is more effective in reducing depression symptoms compared to control conditions. Our publicly released database and interactive dashboard contains over 200 effect sizes from 12 randomized clinical trials testing psilocybin’s impacts on depression and will be updated regularly to keep pace with this rapidly moving field.

*Meaning:* The current evidence suggests promise for psilocybin therapy for depression, though more studies are needed.

## Introduction

Depression is a significant burden on health worldwide, with a growing prevalence and rising economic costs^1^. This high burden has motivated substantial public and private investment in the development of new treatments, including treatment with psychedelics^2^. Psilocybin has seen renewed interest over the last decade driven by emerging evidence of its potential to treat conditions such as depression^3^. Over 130 clinical trials on the therapeutic potential of psilocybin have been initiated in the last two decades, sponsored by over 100 different institutions, and there is estimated to be a potential market size of $10.75 billion for the clinical use of psychedelics by 2027^4^. Thirty-nine of these initiated trials are for treatment-resistant depression or major depressive disorder, for which the U.S. Food and Drug Administration has granted psilocybin Breakthrough Therapy designation to accelerate the approval process.

Given the rapidly growing evidence base, systematic reviews on the topic^5–13^ quickly become outdated, failing to capture the most recent findings. In response, we present a living systematic review on psilocybin treatment for depressive symptoms, ensuring that the latest findings are always reflected in the data synthesis. Our review will be regularly updated; our data, code, and results will be available on our public website: *SYPRES* (Synthesis of Psychedelic Research Studies; <u>sypres.io</u>)^14^. In addition, our continuously maintained database of effect sizes will be released using the Metapsy infrastructure (metapsy.org), and summarized in an interactive online dashboard (metapsy.org/<u>sypres/psilocybin-depression</u>). This dashboard allows scientists, clinicians, and the broader public to retrieve the current evidence on the efficacy of psilocybin for depression. Moreover, it allows individuals to examine the influence of inclusion criteria, analysis choices, and individually-selected subgroups on the variability of study-level effects across this expanding literature. As described below, our initial synthesis of these data suggests that psilocybin shows a significant reduction in depression symptoms compared with control conditions. However, the current evidence base is relatively small and heterogeneous, with limitations that warrant significant caution in the interpretation of these results.

## Methods

This meta-analysis is a pre-registered study under the SYPRES project, and the study protocol is available on PROSPERO (CRD42024584938). Please refer to Supplement 1 for detailed methodological information.

### Eligibility Criteria

We included randomized controlled clinical trials published in English in peer-reviewed journals comparing psilocybin for depressive symptoms with a comparator in adult (>18 years old) populations. Eligible interventions included any dose and formulation (natural or synthetic) of psilocybin or other prodrugs of psilocin intended to produce an alteration of subjective experience in the patient, with or without the conjunctive use of therapy. Eligible comparators included any form of placebo with or without the conjunctive use of therapy, including low doses of the intervention drug and any dose of other psychotropics intended to improve blinding (without known therapeutic efficacy for depression). Eligible comparators also included control for spontaneous improvement via waitlist or usual care. To make our database more comprehensive, we also conducted sensitivity analyses where we expanded these criteria to include reports found in gray literature (Krempien 2023), studies presenting only a mixture of pre– and post-crossover data (Grob 2011), and studies comparing psilocybin with comparators with known therapeutic efficacy (Carhart-Harris 2021). However, these studies were not included in the primary analyses.

### Study search and selection

We searched PubMed, Embase, PsycInfo, Web of Science, Scopus, and the reference lists of systematic reviews retrieved from the searches. Expert research librarians assisted in crafting study search criteria. The final search was conducted on July 1, 2025 (see Supplement 1 for search terms). Study screening and data extraction was independently performed by two reviewers. We assessed risk of bias (RoB) using Cochrane’s Risk of Bias 2.0 tool.

### Effect size calculation

We used each study’s endpoint sample sizes, means, and standard deviations for treatment and control to calculate Hedges’ *g* for each study as the primary effect size measure for continuous outcomes. Our reported effect sizes therefore may differ from study reports using change-from-baseline values (see Supplement 1). For dichotomous outcomes, we calculated risk ratios from raw event data. All analyses, except the three-level model, employed 1 effect size per study.

### Meta-analyses

#### Primary analysis on continuous outcomes

We performed inverse-variance random-effects modeling of standardized mean differences on primary outcomes (see Supplement 1 for selection of outcomes and timepoints for effect sizes). Between-study heterogeneity (*tau*^2^) was calculated using the Restricted Maximum Likelihood (REML) estimator^15^ and the Q-profile method to calculate the confidence interval^16^. We employed the Knapp-Hartung adjustment to calculate the confidence interval for the pooled effect size^17^, which creates a more conservative confidence interval and *p*-value that varies less with changes in heterogeneity variance. Funnel plots and Egger’s test were used to assess small study bias, although the small number of studies limits the precision of these approaches^18^.

#### Three-level CHE meta-analysis and exploratory regression

To assess psilocybin’s effects independent of measurement time point, we applied a three-level meta-analysis model on 30 effect sizes ranging from 1 to 190 days post-dosing generated from the nine studies included in our primary analysis (see Supplement 1). These effect sizes were limited to assessments on the primary instrument occurring at least 1 day after dosing and before any crossover between groups. Variance-covariance matrices of each study with two or more effect sizes were estimated using a within-study correlation coefficient of 0.6, creating a “correlated and hierarchical effects” (CHE) model, which is typically a good approximation for datasets with unknown and/or complex dependence structures^19^. Cluster-robust variance estimation was used to guard against potential model misspecification. Heterogeneity was calculated using the REML estimator with parametric bootstrapping (5,000 iterations) used to generate confidence intervals. To examine the consistency of effects over time, we additionally performed a meta-regression using the three-level CHE model by adding time since the final dose (in days) as a continuous predictor.

#### Meta-analysis on dichotomous outcomes

We also evaluated the dichotomous response and remission outcomes reported by five studies (Ross 2016, Griffiths 2016, Goodwin 2022, Raison 2023, and von Rotz 2023). We used inverse-variance random-effects modeling of risk ratios. Between-study heterogeneity was calculated with the Paule-Mandel (PM) estimator^20^, which is an alternative to REML with a good performance in analyzing dichotomous outcomes. Confidence intervals were again calculated using the Q-profile method for heterogeneity estimates and the Knapp-Hartung adjustment for pooled effect sizes.

#### Subgroup-specific and sensitivity analyses

To assess the robustness of our primary findings and explore potential sources of heterogeneity, we conducted a series of five subgroup-specific analyses for continuous outcomes. These analyses allowed us to evaluate the consistency of treatment effects across different study characteristics and methodological approaches:

1. Depression as primary diagnosis: To assess whether participant diagnosis impacted treatment effects, we conducted a subgroup analysis limited to studies where participants had depression as their primary diagnosis (excluding Griffiths 2016, Ross 2016, and Rieser 2025).
2. Exclude open-label waitlist control: We evaluated the impact of excluding open-label waitlist control studies (Davis, 2021; Rosenblat, 2024).
3. Exclude high RoB: To assess the impact of study quality on outcomes, we excluded studies rated as having high risk of bias (Griffiths 2016 and Rosenblat 2016).
4. Parallel design: We conducted a subgroup-specific analysis that only included parallel group studies (Goodwin 2022, Raison 2023, von Rotz 2023, Back 2024, and Rieser 2025).
5. Crossover design: We conducted an analysis that only included crossover design studies (Griffiths 2016, Ross 2016, Davis 2021, and Rosenblat 2024).

The following sensitivity analyses were also performed:

1. Expanded inclusion criteria: We conducted an analysis with expanded eligibility criteria that incorporated all studies from the primary model plus three additional studies that were excluded from primary analyses (see *Eligibility Criteria*).
2. Excluding outliers: We repeated our primary meta-analysis on continuous outcomes after removing statistical outlier studies (e.g. whose effect size confidence intervals do not overlap with the confidence interval of the pooled effect; Davis 2021).
3. Fixed effects model: We ran fixed-effects models as sensitivity analyses to compare to random effects models. Fixed effects models assume that the between-study variance (*tau*^2^) is 0, such that all studies share a common true effect size. For our continuous model, we used a standard inverse-variance weighting fixed-effects model on standardized mean differences (Hedges’ *g*). For our dichotomous model, we used a standard inverse-variance weighting fixed-effects model on the log risk ratio.
4. Alternate dosing in Goodwin 2022: Given that Goodwin 2022 employed a three-arm design comparing psilocybin at 25 mg and 10 mg doses against a 1 mg control, we conducted a subgroup-specific analysis substituting the 10 mg intervention arm for the 25 mg arm used in our primary analysis.
5. Clinician-rated outcomes: To examine whether the method of assessment influenced observed effects, we conducted a sensitivity analysis that only included clinician-administered depression assessments. MADRS was chosen as the preferred clinician-administered instrument for this analysis, followed by the GRID-HAM-D. Studies included were: Goodwin 2022, Raison 2023, von Rotz 2023, Back 2024, and Rosenblat 2024 using the MADRS; and Griffiths 2016 and Davis 2021 using the GRID-HAM-D.
6. Self-report outcomes: Similarly, we conducted a sensitivity analysis that only included studies reporting self-report depression measures, regardless of whether these were the primary outcome measures. BDI was the preferred self-report instrument which was reported in all self-report studies. Studies included were: Ross 2016, Griffiths 2016, Davis 2021, von Rotz 2023, and Rieser 2025.

#### Software, data, and code availability

Literature screening and data extraction was performed using Covidence. Meta-analyses were conducted using R (4.4.1) in RStudio (2024.04.2+764) with metapsyTools^21^, a package of helper functions for Metapsy that uses meta^22^, metafor^23^, and dmetar^24^ functions. The living database used for this analysis can be accessed through Metapsy (docs.metapsy.org/<u>databases/depression-psiloctr/</u>) or downloaded from our website, where code for all analyses is also available (<u>sypres.io</u>).

## Results

We identified 5,069 reports from our searches. Of these, 82 passed initial title and abstract screening and were reviewed as full-text articles. Nine studies^25–33^ met inclusion criteria for our primary model. An additional three studies^34–36^ were included in the database and our sensitivity analyses on expanded inclusion criteria. One pre-printed study^37^ will be added to the review and database once it is published in a peer-reviewed journal. Complete information about study identification and screening is included in **Figure 1**. Our database consists of 201 effect sizes generated from 12 studies, covering multiple depression instruments, outcome types, and time-points (from baseline to over 6 months). This database has been publicly released^38^ and serves as the basis of our analyses that follow. All studies included a psychotherapy or psychological support component (see Supplement 1). See **Table 1** for detailed study characteristics.

**Figure 1:**
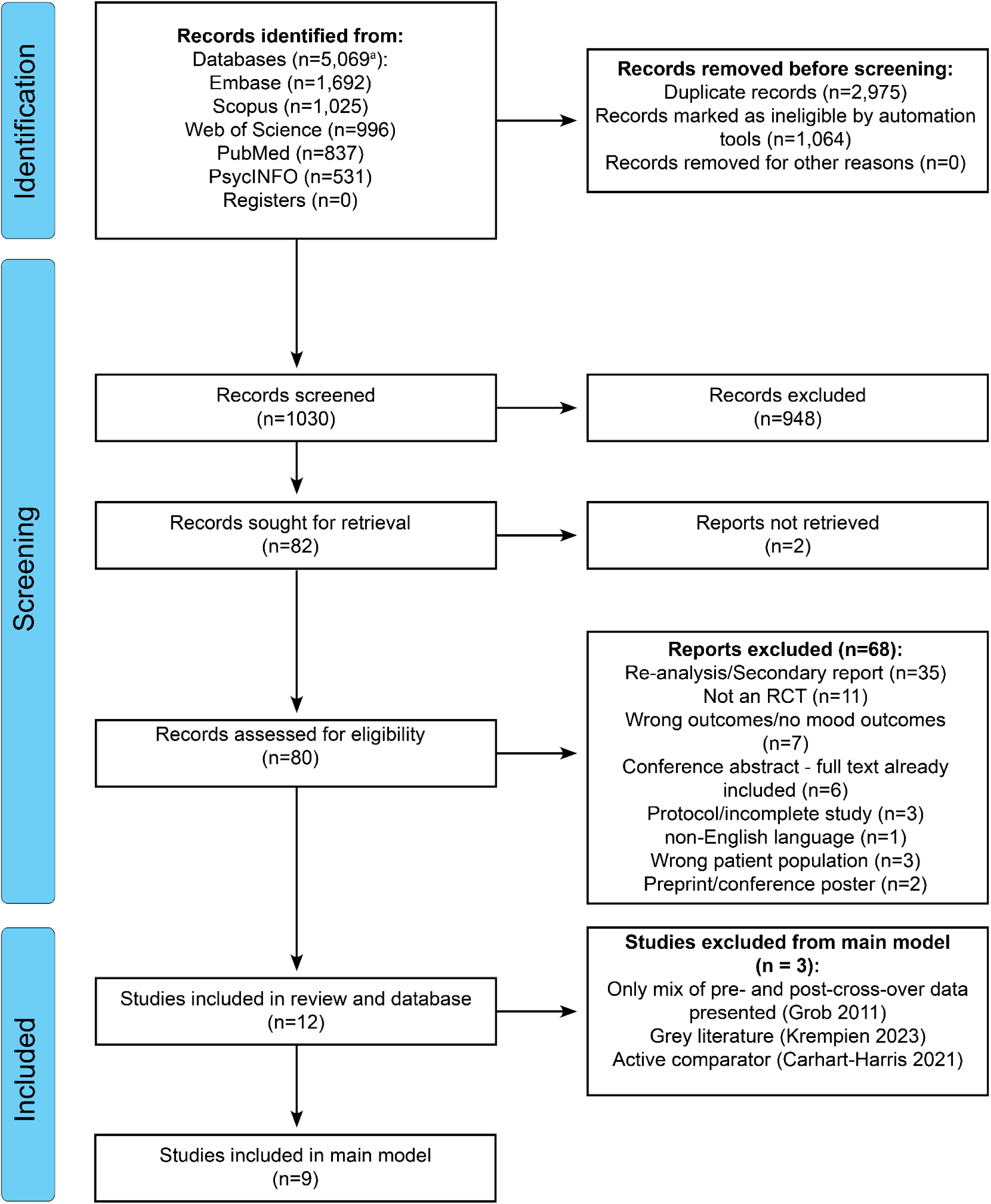
PRISMA 2020 flow diagram. ^a^Includes 12 records that were merged with their parent studies from a total of 5,081 original records; see Supplement 1 for more details.

**Table 1:** Summary of RCTs on psilocybin for depressive symptoms. N given is the number of participants randomized. Study endpoints are the reported primary endpoints for each study and given in weeks since the final dose. Summary statistics across studies (blue rows) were calculated as weighted averages. Overall mean age was calculated using the median age from Grob 2011. ^a^N for these studies reflect the number of participants in the high and low dose groups. Goodwin 2022 and Krempien 2023 contained 75 and 18 participants in the medium dose arms, respectively, bringing the total N for the database to 711. These arms are included in the database, but not used in the present analyses, except the Goodwin 2022 medium dose arm in the alternate dosing for Goodwin et al. sensitivity analysis. US = United States; UK = United Kingdom; CH = Switzerland; CA = Canada; Multi = multi-site. Goodwin 2022 was conducted at sites across the US, CA, UK, and European Union. MDD = major depressive disorder; TRD = treatment-resistant depression; AUD = alcohol use disorder.

| Study ID | N | Country | Age (SD) | Female (%) | Psychedelic use (%) | Patient diagnosis | Intervention | Control | Primary Endpoint |
| --- | --- | --- | --- | --- | --- | --- | --- | --- | --- |
| *Primary meta-analysis* |  |  |  |  |  |  |  |  |  |
| Griffiths 2016 | 56 | US | 56.3 (10.5) | 49 | 45 | Advanced cancer | Single high dose psilocybin (22 or 30 mg/70kg) | Single low dose psilocybin (1 or 3 mg/70kg) | 5 weeks |
| Ross 2016 | 31 | US | 56.3 (12.9) | 62 | 55 | Advanced cancer | Single dose psilocybin (0.3 mg/kg) | Single dose niacin (250 mg) | 7 weeks |
| Davis 2021 | 27 | US | 39.8 (12.2) | 67 | 25 | MDD | Two doses psilocybin (20 mg/70kg; 30 mg/70kg) | Waitlist | 4 weeks |
| Goodwin 2022 | 158<sup>a</sup> | Multi | 39.8 (12.2) | 52 | 6 | TRD | Single high dose psilocybin (25 mg) | Single low dose psilocybin (1 mg) | 3 weeks |
| Raison 2023 | 104 | US | 41.1 (11.3) | 50 | 22 | MDD | Single dose psilocybin (25 mg) | Single dose niacin (100 mg) | 6 weeks |
| von Rotz 2023 | 52 | CH | 36.8 (10.4) | 63 | 31 | MDD | Single dose psilocybin (0.215 mg/kg) | Single dose placebo (mannitol) | 2 weeks |
| Back 2024 | 30 | US | 38 (n/a) | 50 | 7 | MDD | Single dose psilocybin (25 mg) | Single dose niacin (100 mg) | 4 weeks |
| Rosenblat 2024 | 31 | CA | 44.4 (13.7) | 39 | 19 | TRD | Single dose psilocybin (25 mg) | Waitlist | 2 weeks |
| Rieser 2025 | 40 | CH | 37.2 (11.9) | 38 | 51 | AUD | Single dose psilocybin (25 mg) | Single dose placebo (mannitol) | 4 weeks |
| **Total N (primary)** | **529** | - | - | - | - | - | - | - | - |
| **Mean (primary)** | **59** | - | **36.7** | **46** | **24** | - | - | - | **4.1 weeks** |
| *Expanded meta-analysis* |  |  |  |  |  |  |  |  |  |
| Grob 2011 | 12 | US | 36-58 (n/a) | 92 | 67 | Advanced cancer | Single dose psilocybin (0.2 mg/kg) | Single dose niacin (250 mg) | 2 weeks |
| Carhart-Harris 2021 | 59 | UK | 41.2 (10.8) | 34 | 27 | MDD | Two doses psilocybin (25 mg) three weeks apart | Daily escitalopram (10-20 mg) for 6 weeks | 3 weeks |
| Krempien 2023 | 18<sup>a</sup> | US | NA | NA | NA | MDD | Single dose deuterated psilocybin derivative (16 mg) | Single dose placebo (unknown) | 3 weeks |
| **Total N (overall)** | **618** | - | - | - | - | - | - | - | - |
| **Mean (overall)** | **52** | - | **36.2** | **44** | **24** | - | - | - | **3.9 weeks** |

### Risk of bias ratings variable across studies

Overall, of the 12 studies in the database, three studies were determined to have a high risk of bias, five studies had some concerns, and three studies were deemed to have an overall low risk of bias (**Table 2**). Rosenblat 2024 received some concerns for randomization due to the significant imbalance in baseline depression severity between the two arms. This imbalance favors the comparator in our effect size calculation for this study.

**Table 2:** Additional study characteristics and risk of bias assessments using Cochrane’s Risk of Bias 2.0 tool. Randomization = bias due to randomization process; Deviations = bias due to deviations from intended interventions; Missingness = bias due to missing outcome data; Measurement = bias due to measurement of the outcome; Selection = bias due to selection of the reported results; Overall = overall risk of bias; BDI = Beck Depression Inventory; GRID-HAMD = Grid Hamilton Rating Scale for Depression; MADRS = Montgomery-Åsberg Depression Rating Scale; QIDS-SR = Quick Inventory of Depressive Symptomatology–Self-Report.

| Study ID | Design | Scale | Assessor | Risk of Bias Domains |  |  |  |  | Overall |
| --- | --- | --- | --- | --- | --- | --- | --- | --- | --- |
|  |  |  |  | Randomization | Deviations | Missingness | Measurement | Selection |  |
| Primary meta-analysis |  |  |  |  |  |  |  |  |  |
| Griffiths 2016 | Double-blind placebo-controlled | GRID-HAMD | Non-independent study staff | Low | Low | Low | Some concerns | High | High |
| Ross 2016 | Double-blind placebo-controlled | BDI | Self-report | Low | Low | Low | Some concerns | Some concerns | Some concerns |
| Davis 2021 | Open-label waitlist-controlled | GRID-HAMD | Independent study staff | Low | Some concerns | Some concerns | Low | Low | Some concerns |
| Goodwin 2022 | Double-blind placebo-controlled | MADRS | Third-party | Low | Low | Low | Low | Low | Low |
| Raison 2023 | Double-blind placebo-controlled | MADRS | Independent study staff | Low | Low | Low | Low | Low | Low |
| von Rotz 2023 | Double-blind placebo-controlled | MADRS | Non-independent study staff | Low | Low | Low | Some concerns | Low | Some concerns |
| Back 2024 | Double-blind placebo-controlled | MADRS | Independent study staff | Low | Low | Low | Low | Low | Low |
| Rosenblat 2024 | Open-label waitlist-controlled | MADRS | Non-independent study staff | Some concerns | Low | Low | High | Low | High |
| Rieser 2025 | Double-blind placebo-controlled | BDI-II | Self-report | Low | Low | Some concerns | Some concerns | Low | Some concerns |
| Expanded meta-analysis |  |  |  |  |  |  |  |  |  |
| Grob 2011 | Double-blind placebo-controlled | BDI | Self-report | Low | Some concerns | Low | Some concerns | High | High |
| Carhart-Harris 2021 | Double-blind placebo-controlled | QIDS-SR | Self-report | Low | Low | Low | Some concerns | Low | Some concerns |
| Krempien 2023 | Double-blind placebo-controlled | MADRS | Unknown | NA | NA | NA | NA | NA | NA |

### Psilocybin treatment significantly reduces depression symptoms compared with control conditions

The analysis on continuous outcomes in the nine studies included in the primary model showed a statistically significant reduction in depression scores after psilocybin treatment compared with control conditions (**Figure 2**; Hedges’ *g* = –0.91 [-1.35; –0.48], *p* = 0.0013, *k* = 9, *n* = 501), with moderate between-study heterogeneity (*tau*^2^ = 0.13 [0.01; 1.48], *I*^2^ = 58.1% [12.2%; 80.0%]). Visual inspection of a funnel plot (**eFigure 1**) revealed limited asymmetry, and an Egger’s test did not find small study effects (*intercept* = –1.73 [-4.54; –1.08], *t* = –1.21, *p* = 0.27).

**Figure 2:**
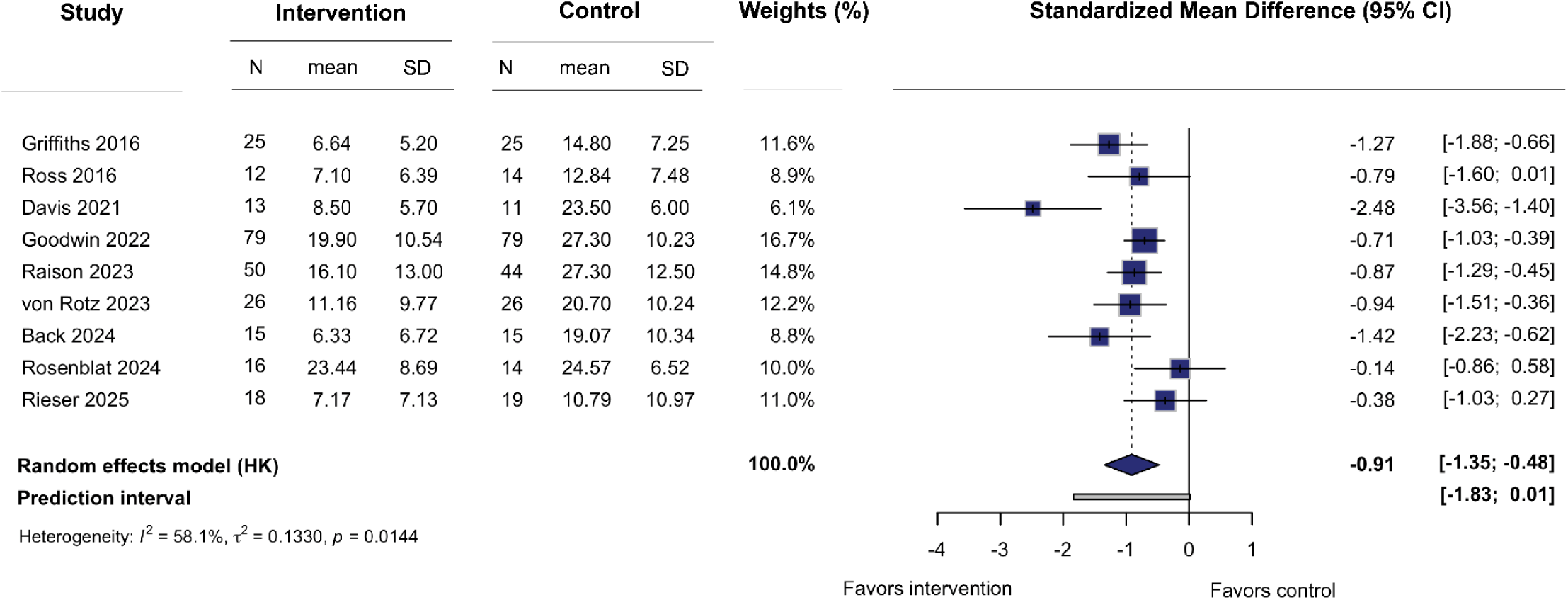
Primary meta-analysis on continuous outcome variables. Boxes represent the standardized mean difference (Hedges’ *g*) for each study, and the lines extending from the box represent the 95% confidence interval around each effect size, while the size of each box is proportional to its weight. The diamond at the bottom represents the pooled effect size (meta-analytic mean). The gray line at the bottom represents the prediction interval of the expected range of true effects in a new study. HK = Knapp-Hartung adjustment.

### Psilocybin’s effects are rapid and consistent over several weeks

Our three-level CHE model revealed an overall significant decrease in depression scores with psilocybin compared to the control conditions (Hedges’ *g* = –0.90 [-1.23; –0.57], *p* < 0.001, *k* = 9, *n* = 529, *tau*^2^ = 0.18 [0.03; 0.47], *I*^2^ = 74% [31%; 88%]), ensuring that our results were not sensitive to the time point studied. Adding time since final dose as a continuous predictor to our model (**eFigure 2**) revealed a significant effect favoring psilocybin immediately following dosing (*intercept* = –0.92 [-1.26; –0.58], *p* < 0.0001) that was stable over time (*slope* = 0.0009 [-0.0023; 0.0041], *p* = 0.57).

### Higher response and remission rates after psilocybin treatment

In addition to differences in symptoms of depression, we found evidence for statistically significant greater treatment response with psilocybin compared with control conditions (**eFigure 3**; *RR* = 2.77 [1.93; 3.97], *p* = 0.001, *k* = 5, *n* = 380), with low between-study heterogeneity (*tau*^2^ = 0.00 [0.00; 0.96]; *I*^2^ = 0.00% [0.00%; 79.20%]). We also found that there were significantly higher remission rates with psilocybin compared with control conditions (**eFigure 4**; *RR* = 4.13 [3.39; 5.04], *p* < 0.001, k = 5, n = 380), with low between-study heterogeneity (*tau*^2^ = 0.00 [0.00; 0.00]; *I*^2^ = 0.00% [0.00%; 79.20%]).

**Figure 3:**
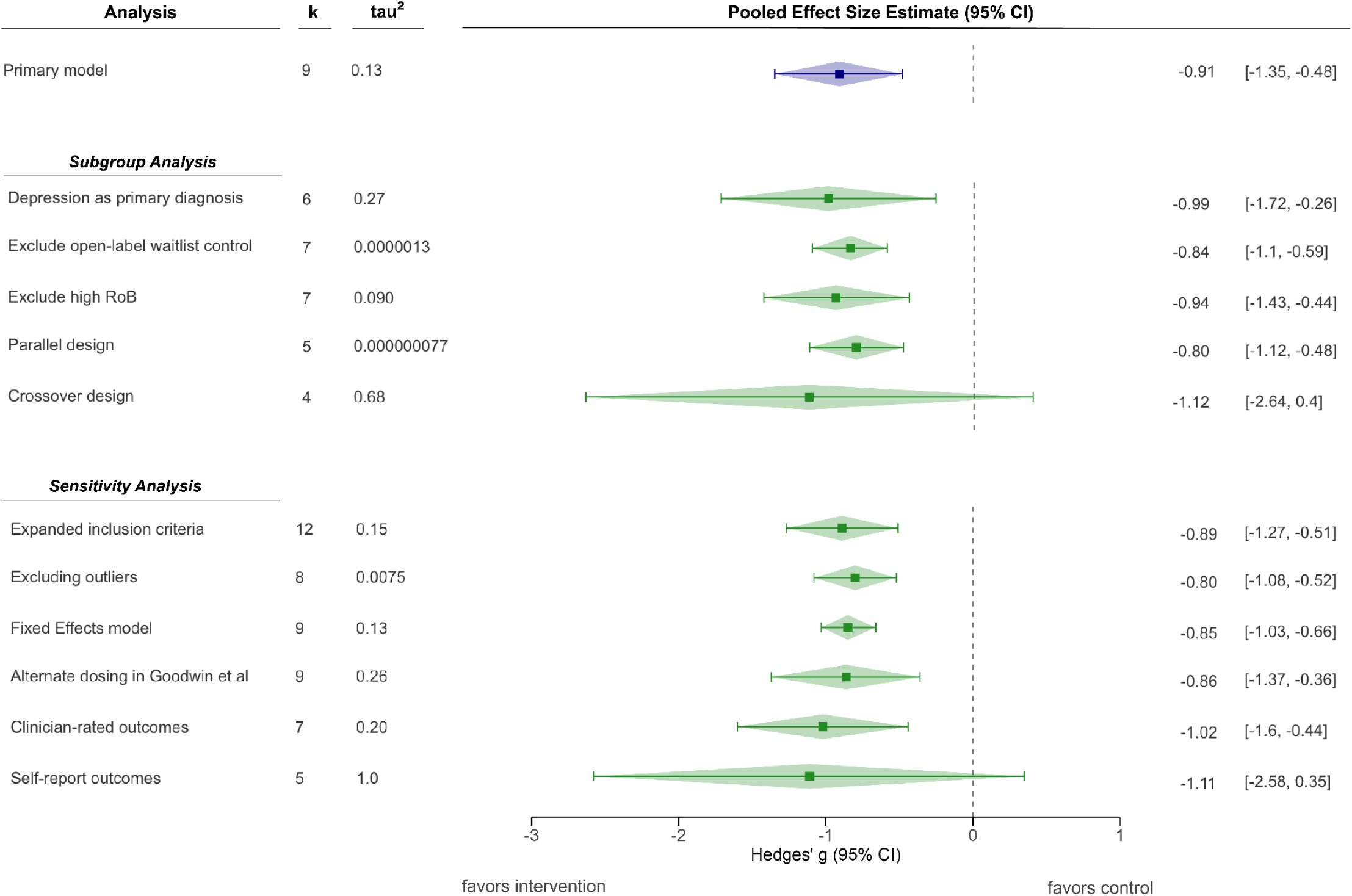
Pooled effect sizes for subgroup-specific and sensitivity analyses. Box and whiskers represent the meta-analytic mean and corresponding 95% confidence intervals for each subgroup-specific analysis. K and *tau*^2^ represent the number of studies included in each analysis and heterogeneity for each analysis, respectively. The pooled effect size from the primary model is presented at the top for comparison purposes.

### Subgroup-specific analyses reveal open-label waitlist control and crossover designs as high sources of heterogeneity

Our subgroup-specific analyses produced results largely in line with our primary model results. Hedges’ *g* values of each subgroup-specific analysis were largely similar to the primary model, with four out of five analyses showing significant results (**Figure 3**). Notably, subgroup-specific analyses excluding either open-label waitlist control studies or crossover studies had substantially lower between-study heterogeneity *(tau*^2^ = 1.3×10^−6^ [0; 0.47] and 7.7×10^−8^ [0.00; 1.05], respectively). Analyzing crossover studies on their own resulted in both higher heterogeneity (*tau*^2^ = 0.68 [0.09; 13.33]) and a non-significant effect of psilocybin (−1.12 [−2.64; 0.40]). It is worth noting, however, that the four studies included here vary in other important factors as well (*k* = 2 open-label waitlist control, *k* = 2 advanced cancer).

### Sensitivity analyses show significant and comparable results to the primary model

We performed a series of sensitivity analyses that largely supported our primary results. First, our model using expanded inclusion criteria showed a significant and comparable effect size (**eFigure 5**; Hedges’ *g* = –0.89 [-1.27; –0.51], *p* <0.001, *k* = 12, *tau*^2^ = 0.15 [0.03; 1.21], *n* = 602). We also ran a model removing statistical outliers (Davis 2021), resulting in a significant effect size comparable to the primary model (Hedges’ *g* = –0.80 [-1.08; –0.52], *p* <0.001, *k* = 8, *tau*^2^ = 0.0075 [0.00; 0.61], *n* =477). Furthermore, we replicated our primary analyses using a fixed effects model on both continuous outcomes (Hedges’ *g* = –0.85 [-1.03; –0.66], *p* < 0.001, k = 9, *n* = 501), response outcomes (*RR* = 2.8 [2.05; 3.83], *p* < 0.001, *k* = 5, *n* = 380), and remission outcomes (*RR* = 4.13 [2.66; 6.42], *p* <0.001, *k* = 5, *n* = 380). In contrast, a model evaluating self-report outcomes yielded insignificant results with substantial heterogeneity (Hedges’ *g* = –1.11 [-2.58; 0.35], *p* = 0.102, *k* = 5, *tau*^2^ = 1.03 [0.22; 13.1], *n* = 190). It should be noted, however, that the five studies included in the self-report analysis vary substantially by treatment population (*k* = 2 MDD, *k* = 2 advanced cancer, and *k* = 1 AUD). All remaining sensitivity analyses showed significant and comparable results (**Figure 3**).

## Discussion

There is an urgent need for new therapies for depression^39,40^. Current evidence, synthesized here, suggests that psilocybin may have therapeutic potential for treating depression symptoms, with a large overall effect size. Compared to placebo, persons who received psilocybin had significantly lower depression scores, higher response and remission rates, with minimal variation in effect sizes by inclusion criteria or analysis choices, and these effects were rapid and consistent over several weeks. However, there was increased heterogeneity associated with study design and variability in risk of bias that should be considered when interpreting these findings. We present these results alongside an open-resource database, codebase, and dashboard – providing a living resource that will be regularly updated as new evidence emerges.

Our work improves upon prior literature in several significant ways. First, while our results are consistent with prior meta-analyses, previous systematic reviews on the topic have quickly become outdated^5–13^. Second, in addition to our commitment to updating these results as a living review, our data and code are publicly accessible – maximizing transparency and reproducibility. Third, we also included numerous sensitivity analyses, which confirmed that our results are not driven by specific inclusion criteria or model parameters. Fourth, our subgroup-specific analyses identified potential sources of heterogeneity in the literature, including open-label waitlist-control studies and crossover study designs. Finally, our hierarchical effects meta-analysis and meta-regression provide new results suggesting that psilocybin’s effects over time are rapid and consistent over several weeks.

However, there remain substantial challenges in synthesizing evidence from currently available data^41^. Most psilocybin for depression trials to date have been small, with substantial variability in design and methods. In addition, study populations were often carefully selected and may not be representative of the general population that might receive treatment if psilocybin receives regulatory approval. As such, RCTs in the field often struggle with external validity^42–47^. Hence, future studies with larger treatment groups and expanded inclusion criteria will aid the generalizability of these findings.

Issues such as functional unblinding and expectancy effects also complicate interpretation. Double-blinding in psychedelic-assisted psychotherapy has been the topic of extensive discourse in the field^48–51^. The psychoactive effects of psilocybin make functional unblinding likely, and studies that have tested functional unblinding find that almost all participants and/or blinded study staff correctly guess group assignments^25,26,31,33^. In light of this, a recent meta-analysis comparing effect sizes of psilocybin treatment with open-label selective serotonin-reuptake inhibitor (SSRI) trials finds the two therapies are nearly identical in their efficacy for depression relative to within-arm control^52^. In addition to increased expectancy in arms receiving psilocybin treatment, there may be a substantial under-performance in the placebo arm, as evidenced by an underperformance of placebo arms in psychedelic trials compared with placebo arms in trials of escitalopram (an SSRI)^51–53^. Given this potential for inflated effect sizes in psilocybin trials, our large pooled effect size should be interpreted with caution. Furthermore, the power of our results is limited given the small number of studies that met our eligibility criteria and longer duration studies are needed to determine the durability of psilocybin’s effects beyond a few months. Finally, we found either some concerns or high risk of bias in most studies, even without fully accounting for potential bias from functional unblinding or expectancy effects.

## Conclusions

Within the context of these important limitations, our results suggest that psilocybin may be a promising treatment for symptoms of depression, though larger studies with expanded patient populations and rigorous methods are needed. As more RCTs are published, we will regularly update our SYPRES website and dashboard in a reproducible and transparent manner. As part of our SYPRES initiative, we will also conduct a series of future meta-analyses on other psychedelic therapies, including MDMA for PTSD and psilocybin for anxiety^14^. This living systematic review and open science resource will provide a valuable and transparent resource for researchers, clinicians, policymakers, and the public.

## Article information

### Author contributions

SPS and BLS are joint first authors. JCS and TDS are joint senior authors.

Concept and design: All authors.

Acquisition, analysis, or interpretation of data: All authors.

Drafting of the manuscript: SPS, BLS, JCS, and TDS.

Critical revision of the manuscript for important intellectual content: All authors.

Statistical analysis: SPS, BLS, MJ, SNV, JCS, TDS.

Obtained funding: RHD and TDS.

Administrative, technical, or material support: PM and MH.

Supervision: JCS and TDS.

### Additional contributions

We thank Jen Lege-Matsuura and Kristina McShea for their assistance with generating database search terms. We thank Nathan Sepeda, Robin von Rotz, Joshua Rosenblat, Nathalie Rieser, Robin Carhart-Harris, Balázs Szigeti, David Erritzoe, Amir Inamdar, and Sasha Kulikova for providing data or other clarifications on studies included in this review.

## Data Availability

The living database used for this analysis can be accessed through Metapsy (docs.metapsy.org/databases/depression-psiloctr/) or downloaded from our website, where code for all analyses is also available sypres.io).

https://sypres.io

https://www.metapsy.org/sypres/psilocybin-depression

https://docs.metapsy.org/databases/depression-psiloctr/

