## Supplement 1 for "Psilocybin treatment for symptoms of depression: a living systematic review, meta-analysis, and data resource"

**Supplementary Figures** 2
**Supplementary Methods** 5
Search and selection 5
Data extraction 6
Risk of bias assessment 6
Study harmonization 8
Selection of effect sizes 9
**Database description** 11
**Search Terms** 13
Pubmed 13
Embase 14
PsycInfo 14
Web of Science 15
Scopus 15
**Psychotherapy or Psychological Support** 15
**Excluded Reports** 17
**Merged and Excluded Secondary Reports** 23
**Record of Contacting Study Authors** 25
**Risk of Bias Assessment Standard Operating Procedure** 33
**Overall Workflow** 33
**Domain 1 - Randomization** 33
**Domain 2 - Deviations** 34
**Domain 3 - Missing Data/Dropout** 35
**Domain 4 - Measurements** 35
**Domain 5 - Pre-registration** 36
**Supplementary References** 36

### Supplementary Figures

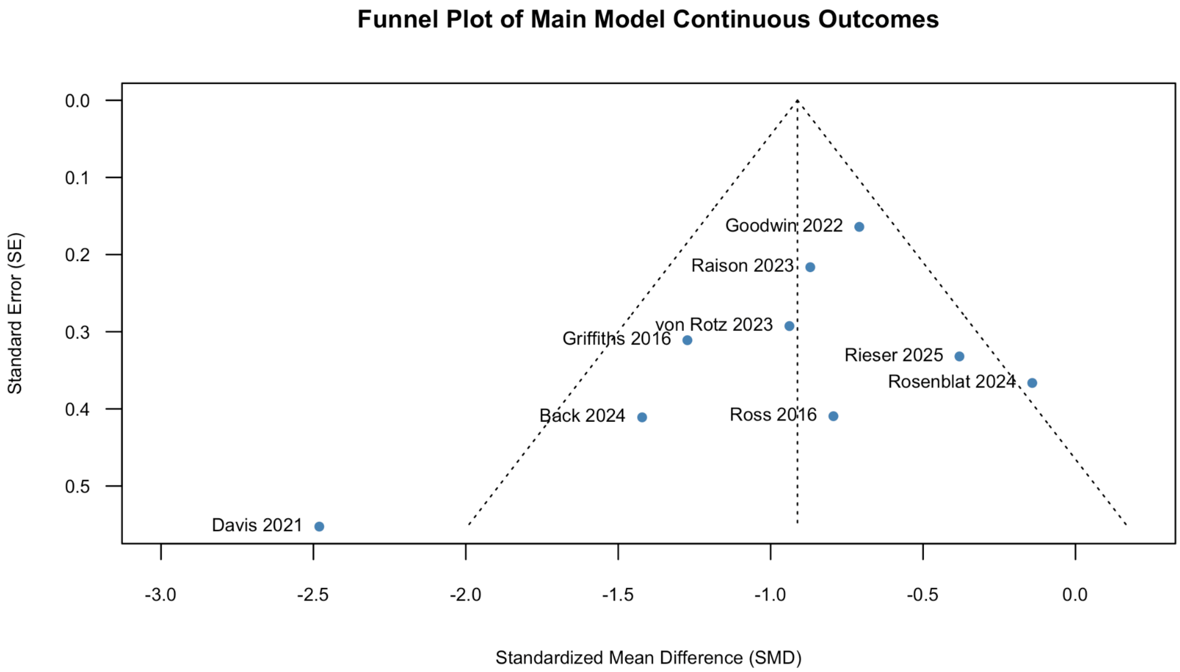

**eFigure 1**: Funnel plot of 9 studies in primary meta-analytic model.

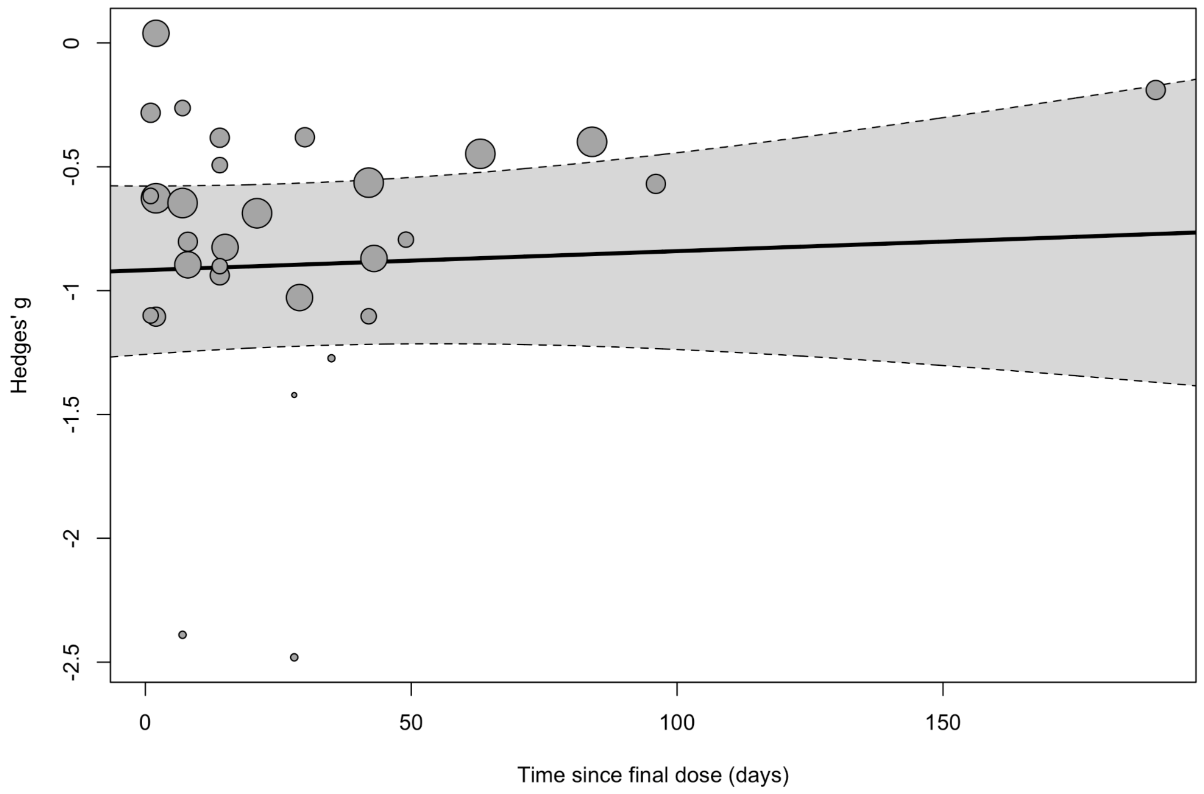

**eFigure 2**: Meta-regression of time since final dose on the three-level CHE model.

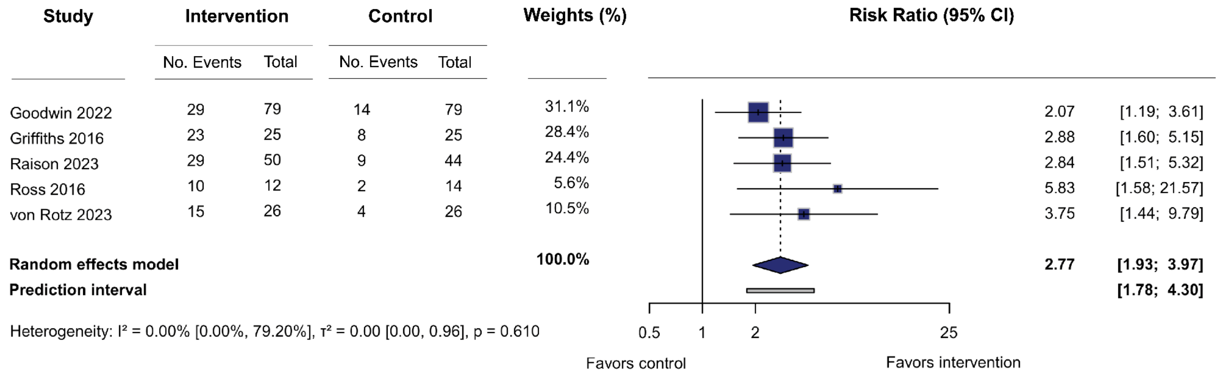

**eFigure 3**: Meta-analysis on response to treatment, indicating improvement in depression symptoms per clinically significant change from baseline score (as defined by each study). Boxes represent the standardized mean difference (Hedges' *g*) for each study, and the lines extending from the box represent the 95% confidence interval around each effect size, while the size of each box is proportional to its weight. The diamond at the bottom represents the pooled effect size (meta-analytic mean). The gray line at the bottom represents the prediction interval of the expected range of true effects in a new study.

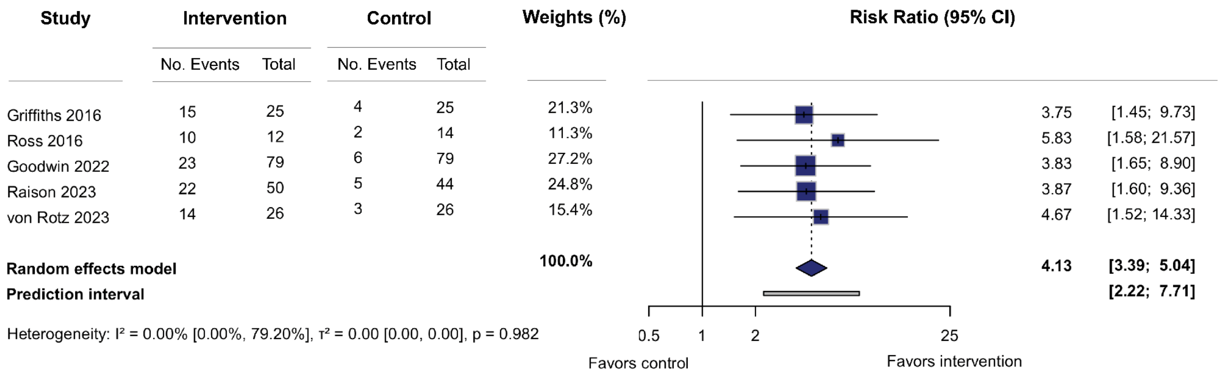

**eFigure 4**: Meta-analysis on remission rates. Boxes represent the standardized mean difference (Hedges' *g*) for each study, and the lines extending from the box represent the 95% confidence interval around each effect size, while the size of each box is proportional to its weight. The diamond at the bottom represents the pooled effect size (meta-analytic mean). The gray line at the bottom represents the prediction interval of the expected range of true effects in a new study.

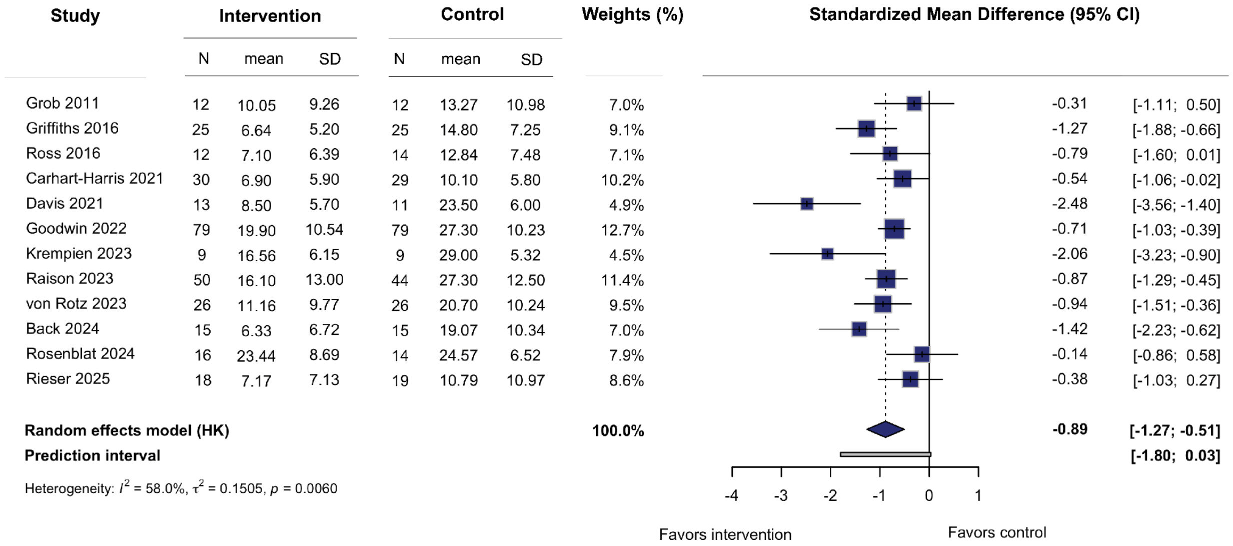

**eFigure 5**: Forest plot of 12 studies in the expanded model. Boxes represent the standardized mean difference (Hedges' *g*) for each study, and the lines extending from the box represent the 95% confidence interval around each effect size, while the size of each box is proportional to its weight. The diamond at the bottom represents the pooled effect size (meta-analytic mean). The gray line at the bottom represents the prediction interval of the expected range of true effects in a new study. HK = Knapp-Hartung adjustment.

### Supplementary Methods

#### Search and selection

We searched PubMed, Embase, PsycInfo, Web of Science, and Scopus. Additionally, we searched reference lists of prior systematic reviews identified from our primary search. Our search syntax combined MeSH and text terms under the PICO (Population, Intervention, Comparator, Outcome) framework to identify randomized controlled trials on the effects of psilocybin on mood and depression outcomes (see *Search Terms* for a complete list of search syntax for each database). The search syntax was developed in collaboration with research librarians with expertise in systematic reviews. The initial database searches occurred from September 24, 2024 through October 11, 2024. The final search was conducted on July 1, 2025. The publication date range for included studies was from the database start range to the final search date. Updated searches will be periodically conducted given the living nature of this systematic review, with updated results published on our SYPRES website.

Search results were saved and loaded into Covidence, a web-based collaboration software platform that streamlines the production of systematic and other literature reviews. Covidence automatically filtered out duplicates and auto-marked non-RCTs as ineligible. Two team members (SPS & BLS) performed screening of articles based on predefined exclusion and inclusion criteria (see *Eligibility Criteria* in the main text). In screening, we evaluated the title and abstract of all references returned by our search. If both reviewers voted to include the reference in full text review, then it was moved forward to the next phase of evaluation. Conflicts were resolved via a consensus discussion before the next phase. In the next phase, both reviewers independently evaluated the full text of the article. Disagreements on either inclusion or exclusion (or on the reason for exclusion) were resolved via a consensus discussion or discussion with a third reviewer (TDS or JCS). Reasons for excluding a report were determined on a 5-level hierarchical basis. First, if the report was not an RCT or presented in a language other than English, it was excluded. Second, if neither of these applied, but the report represented a published protocol of an upcoming RCT or initial results from an incomplete RCT, it was excluded. Third, if neither of the first two levels applied, then if the report was a preprint or conference abstract of a study where future peer-reviewed results were anticipated but not yet published, it was excluded. Fourth, if the report did not present any mood outcomes or if it was the wrong patient population (e.g. healthy individuals or pediatric patients) then it was excluded. Fifth and finally, if none of the previous levels applied, then any of the following reasons could qualify as an exclusion reason: the report was a conference abstract of a study where the full-text was later published, or a secondary report of an already included original study. If none of these reasons applied, the report was included. If a secondary report contained information possibly relevant to current or future versions of our database, we included it and merged it with the parent study. See *Excluded Reports* for the full list of reports excluded during full-text review and their reasons. The two reviewers agreed on 95% and 96% of title/abstract screening and full-text review decisions, respectively. Complete details of the study selection process including reasons for exclusion are provided in **Figure 1**.

#### Data extraction

Data was independently extracted from included studies by two team members (BLS & AL or SPS) using Covidence. Resources used to extract the data included the primary paper, supplemental data, clinical trial registries, and protocol details. Follow-up papers published on the same clinical trial that either added additional time points or more mood-related outcomes to the original paper were grouped with the original paper and extracted as one study to avoid duplication.

Extracted data included identification information, such as author details, setting, and time frame of clinical trial; methods information, such as study design, randomization and recruitment methods, blinding, assessment and therapy details, outcome details, the primary time-point and outcome, and statistical analyses used; population information, such as inclusion and exclusion criteria, withdrawal details, and baseline characteristics; intervention information, such as allocation to groups, dosage information, and frequency of drug administration; and outcomes and results information. For outcomes and results, we extracted all relevant mood outcomes at every timepoint provided in the paper. We extracted both dichotomous and continuous outcome measures in the format provided by the paper or supplementary data, prioritizing raw data over effect sizes. The dichotomous outcomes of response and remission were defined internally within each study and not by the authors of this review.

If information pertinent to the meta-analysis were not available, authors were contacted via e-mail. If desired data were available only in figures and not reported in the article text or successfully obtained via author contact, we used the WebPlotDigitizer tool[1] to extract results data.

Disagreements in data extraction were resolved via consensus discussion or by a third reviewer (SPS). For WebPlotDigitizer data, one team member (BLS) used the tool to extract results, while another team member (AL) checked the results. In case of disagreement, the second reviewer replicated the data extraction using the WebPlotDigitizer tool, and remaining disagreements were resolved by a consensus discussion.

#### Risk of bias assessment

We assessed study bias using Cochrane's Risk of Bias (RoB) 2.0 tool[2,3], which is the standard approach for bias assessment in randomized controlled trials. We focused our assessment specifically on primary outcome variables. In studies with multiple primary outcomes or where the primary outcome was not a depression measure, we evaluated the outcome selected for our primary meta-analysis (see *Selection of Effect Sizes* section in supplement). Furthermore, we limited our bias assessment to pre-crossover study periods, as our analyses here were restricted to pre-crossover data. If information pertinent to assessment of risk of bias was not available, authors were contacted via email. Risk of bias was not assessed on gray literature (Krempien 2023).

The RoB 2.0 tool examines five potential bias sources: randomization procedures, deviations from the intended protocol, missing outcome data, outcome measurement, and selective reporting. Each domain contains several questions rated on a four-level scale ranging from ‘yes’ to ‘no’, in addition to an option for insufficient information. Domain-level and overall study bias ratings are classified as low, medium, or high according to a predetermined algorithm[3]. While this algorithm guides assessment, evaluators may justifiably override automated bias determinations when specific concerns warrant greater or lesser emphasis than the algorithm suggests. Two team members (SPS & BLS) independently conducted the bias assessment. Through collaborative discussion and re-examination, at times with additional team members (SMN & JCS), consensus was ultimately achieved. **Table 2** presents the final assessments; Supplement 2 details individual items.

We used the handbook associated with the RoB 2.0 Tool to interpret the signaling questions, but some of the questions had particular nuances in the context of psychedelic-assisted therapy, which were discussed by team members before conducting RoB assessments to ensure consistent interpretations. See *Risk of Bias Assessment Standard Operating Procedure* for the full risk of bias assessment procedure that was used in conjunction with the Cochrane tool guidelines and handbooks to answer signaling questions.

An important consideration of psychedelic-assisted psychotherapy trials is the high potential for functional unblinding due to the unique psychoactive effects of psilocybin. We took this into account when answering risk of bias questions in both Domain 2, focusing on protocol deviations, and Domain 4, focusing on the outcome measurement. Signaling questions 2.1 and 2.2 ask respectively if participants and carers or people delivering the intervention were aware of participants’ assigned intervention during the trial. Although most of the studies in our meta-analyses were methodologically double-blind studies, due to the psychoactive effects in the psilocybin group, we answered ‘PY’ to this question in most cases. If functional unblinding was measured in either participants and/or study staff, we answered ‘Y’ as all of these studies found evidence for high degrees of functional unblinding. Note that while our answer of ‘Y’ for studies that actually assessed functional unblinding indicates an increased confidence in the answer to the signaling question, in the risk of bias algorithm responses of ‘PY’ and ‘Y’ are weighted equally.

In domain 4, signaling question 4.3 asked if outcome assessors were aware of the intervention received by study participants. In this case, the relevance of functional unblinding depended on who the outcome assessors were. For self-report questionnaires, the outcome assessor was the participant. For clinician-administered questionnaires, outcome assessors were either blinded raters (such as independent study staff or third party raters), or unblinded raters (such as a non-independent study staff who were involved in the administration of therapy or other forms of patient contact). Following the same logic used when answering signaling question 2.1 and 2.2, we answered 4.3 as follows. Self-report questionnaires and unblinded (functionally or otherwise) clinician raters received ‘PY’ or ‘Y’, and clinician raters who were blinded independent study staff or third-party raters received ‘PN’ or ‘N’. Answering ‘PN’ or ‘N’ on 4.3 ends the algorithm and signals a low risk of bias for domain 4. Thus, studies that used blinded independent study-staff or third-party raters to administer clinician assessments received a low risk of bias for this domain. If any other answer is given for 4.3, two additional questions follow. Specifically, 4.4 asks if knowledge of the intervention *could* influence the assessment, to which we always answered ‘Y’, and 4.5 asks if knowledge of the intervention *is likely* to influence the assessment. Here, we considered self-report data to have less risk of bias than explicitly unblinded (open-label) non-independent study-staff rating clinician-administered scales. A non-independent study staff potentially had more bias or stake in the “success” of psilocybin treatment, potentially contributing to bias in the way they surveyed the outcome measure, requiring an answer of ‘PY’ for 4.5. However, there is evidence that self-reports actually have low risk of bias and do not over-inflate the effects of the intervention treatment [4], justifying a response of ‘PN’ for 4.5. For clinician-administered outcomes by other non-independent study staff, a judgement call must be made based on their level of patient contact, potential for functional unblinding, and overall involvement in the study. Domain 4 assessment is summarized in **eTable 1** below:

| Assessor | Domain 4 Result |
|---|---|
| Blinded third-party independent rater | Low |
| Blinded independent study staff | Low |
| Potentially functionally unblinded non-independent study staff | Some concerns or High[a] |
| Explicitly unblinded (open-label) non-independent study staff | High |
| Self | Some concerns |

**eTable 1:** Summary of the relationship between the identity of the assessor and risk of bias rating on domain 4. [a]May be upgraded to ‘High’ depending on the rater’s level of patient contact, potential for functional unblinding, and overall involvement in the study.

#### Study harmonization

To ensure methodological consistency when calculating standardized mean differences (SMDs), we addressed an often overlooked issue in continuous outcome meta-analyses: the discrepancy between studies reporting endpoint data versus those reporting change-from-baseline data[5]. Since SMD calculations require that pooled standard deviations in the denominator reflect comparable variance across studies[6], we implemented a systematic approach to harmonize these measurements.

Four studies (Carhart-Harris 2021, Goodwin 2022, Krempien 2023, and Rosenblat 2024) initially reported only change scores and associated standard deviations. We first contacted all authors to request endpoint means and standard deviations. This approach was successful for three studies (Carhart-Harris 2021, Krempien 2023, and Rosenblat 2024), from which we obtained the necessary endpoint data. For the Goodwin study, where author contact was not successful, we implemented an imputation strategy for the Montgomery–Åsberg Depression Rating Scale (MADRS) endpoint standard deviations. Specifically, we calculated the average endpoint standard deviations from five comparable studies that used the MADRS scale (Carhart-Harris 2021, Raison 2023, von Rotz 2023, Back 2024, Rosenblat 2024). We derived separate average standard deviation for the intervention and control arms, then applied these imputed standard deviations to the respective arms of the Goodwin study. Both the 25 mg and 10 mg arms were treated as 'intervention' arms while the 1 mg arm was treated as control for imputations. For Raison 2023 and Rosenblat 2024, endpoint values were available only for the primary endpoint. When imputing endpoint standard deviations for other timepoints from these studies, we used each study's own primary endpoint standard deviation.

#### Selection of effect sizes

For all models except the three-level correlated and hierarchical effects (CHE) model (see below), only one effect size was used per study. In most cases, this was the Hedges' *g* calculated at that study's primary endpoint, using their own reported primary outcome depression instrument. For studies that did not report a primary depression instrument, or reported multiple, we identified one to serve as primary (**eTable 2**). In these cases, we first prioritized clinician-administered depression instruments, when available, and then instruments used the most in other studies. For studies that did not report a primary endpoint, or reported multiple primary endpoints, we selected the last time point in the post-intervention study period prior to any crossover event or follow-up period (**eTable 3**). For the five studies which reported dichotomous outcomes, we used effect sizes (risk ratios) for the same instrument and timepoint as was used in the primary continuous analysis. For our three-level CHE model on continuous outcomes, we used any effect size extracted from a study that was derived from the selected primary instrument during any time point in the post-intervention study period prior to any crossover event or follow-up period (**eTable 4**). See **eTable 5** for the full list of outcomes and timepoints extracted in each study.

| Study | Reported Primary Instrument(s) | Selected Instrument |
|---|---|---|
| Griffiths 2016 | GRID-HAMD-17; HAM-A | GRID-HAMD-17 |

| | | |
|---|---|---|
| Ross 2016 | HADS Total; HADS-A; HADS-D; STAI Trait; STAI State; BDI | BDI |
| Davis 2021 | GRID-HAMD | GRID-HAMD |
| Goodwin 2022 | MADRS | MADRS |
| Raison 2023 | MADRS | MADRS |
| von Rotz 2023 | MADRS; BDI | MADRS |
| Back 2024 | MADRS | MADRS |
| Rosenblat 2024 | MADRS | MADRS |
| Rieser 2025 | Abstinence; Mean alcohol use (Timeline Followback Method) | BDI |
| Grob 2011 | STAI; BDI; POMS | BDI |
| Carhart-Harris 2021 | QIDS-SR-16 | QIDS-SR-16 |
| Krempien 2023 | MADRS | MADRS |

**eTable 2**: Primary instruments reported by each study along with the instrument used for our analyses.

| Study | Reported Primary Endpoint(s) | Selected Endpoint |
|---|---|---|
| Griffiths 2016 | 5 weeks after session 1 | 5 weeks after session 1 |
| Ross 2016 | 1-day post-Dose 1; 2 weeks post-Dose 1; 6 weeks post-Dose 1; 7 weeks post-Dose 1 (1 day pre-dose 2) | 7 weeks post-dose 1 (1 day pre-dose 2) |
| Davis 2021 | Week 5; Week 8 | Week 8 |
| Goodwin 2022 | Week 3 | Week 3 |
| Raison 2023 | Day 43 | Day 43 |
| von Rotz 2023 | Day 14 | Day 14 |
| Back 2024 | Day 28 | Day 28 |

| | | |
|---|---|---|
| Rosenblat 2024 | Day 14 | Day 14 |
| Rieser 2025 | Day 30 | Day 30 |
| Grob 2011 | 1 day before experimental session; 1 day after experimental session; 2 weeks after experimental session; 1 month; 2 months; 3 months; 4 months; 5 months; 6 months | 2 weeks after experimental session |
| Carhart-Harris 2021 | Week 6 | Week 6 |
| Krempien 2023 | Day 21 | Day 21 |

**eTable 3**: Primary endpoints reported by each study along with the endpoint selected for our analyses.

| Study | Effect sizes (N) | Instrument | Time points |
|---|---|---|---|
| Griffiths 2016 | 1 | GRID-HAM-D | 5 weeks |
| Ross 2016 | 4 | BDI | 1 day, 2, 6, and 7 weeks |
| Davis 2021 | 2 | GRID-HAM-D | 1 and 4 weeks |
| Goodwin 2022 | 6 | MADRS | 2 days, 1, 3, 6, 9, and 12 weeks |
| Raison 2023 | 5 | MADRS | 2, 8, 15, 29, and 43 days |
| von Rotz 2023 | 3 | MADRS | 2, 8 and 14 days |
| Back 2024 | 1 | MADRS | 4 weeks |
| Rosenblat 2024 | 3 | MADRS | 1, 7, and 14 days |
| Rieser 2025 | 5 | BDI | 1, 14, 30, 96, and 190 days |

**eTable 4**: List of the 30 effect sizes selected for the three-level CHE model. Time points are given in reference to the final dosing day.

### Database description

Our publicly released database contains 201 total effect sizes generated from 12 studies (**eTable 5**). These effect sizes encompass all depression outcomes and all timepoints reported by arm in each study. Effect sizes are generated from five different outcome types (**eTable 6**).

Full, up-to-date documentation on the database, including descriptions of all variables, can be found here: https://docs.metapsy.org/databases/depression-psiloctr/.

| Study | Effect sizes (N) | Instruments | Timepoints |
|---|---|---|---|
| Griffiths 2016 | 15 | BDI, GRID-HAM-D, HADS-D | 5 weeks after session 1, 5 weeks after session 2, 6 months |
| Ross 2016 | 26 | BDI, HADS-D | 1 day pre-dose 1, 1 day post-dose 1, 2 weeks post-dose 1, 6 weeks post-dose 1, 7 weeks post-dose 1 and 1 day pre-dose 2, 6 weeks post-dose 2, 26 weeks post-dose 2 |
| Davis 2021 | 7 | BDI, QIDS-SR, GRID-HAM-D, PHQ-9 | Week 5, Week 8 |
| Goodwin 2022 | 81 | QIDS-SR, MADRS | Week 3, Week 12, Day 2, Week 1, Week 6, Week 9, Week 3-12 Sustained Response |
| Raison 2023 | 22 | MADRS, SMDDS | Day 2, Day 8, Day 15, Day 29, Day 43, Days 8-43 Sustained Response |
| von Rotz 2023 | 14 | BDI, MADRS | Day -1, Day 0, Day 2, Day 8, Day 14 |
| Back 2024 | 2 | MADRS | Day -8, Day 28 |
| Rosenblat 2024 | 6 | MADRS | Day 1, Day 7, Day 14 |
| Rieser 2025 | 6 | BDI | Day -14, Day 1, Day 14, Day 30, Day 96, Day 190 |
| Grob 2011 | 3 | BDI | 1 day before experimental session, 1 day after experimental session, 2 weeks after experimental session |
| Carhart-Harris 2021 | 16 | QIDS-SR, HAM-D-17, BDI-1A, MADRS | Week 6 |
| Krempien 2023 | 3 | MADRS | Day 21 |

**eTable 5**: List of effect sizes compiled in the full database released through Metapsy. Timepoints here are verbatim references to the text used in the study reports.

| Outcome type | Effect size | Inputs |
|---|---|---|
| msd | Hedges' *g* | 1. Endpoint N's<br>2. Endpoint means<br>3. Endpoint standard deviations |
| imsd | Hedges' *g* | 1. Endpoint N's<br>2. Endpoint means calculated by adding change score means to baseline means<br>3. Imputed endpoint standard deviations (see *Study harmonization* for details) |
| change | Hedges' *g* | 1. Endpoint N's<br>2. Endpoint means calculated by adding change score means to baseline means<br>3. Change score standard deviations[a] |
| response | Log Risk Ratio | 1. N responders at endpoint<br>2. N participants at endpoint |
| remission | Log Risk Ratio | 1. N remitters at endpoint<br>2. N participants at endpoint |

**eTable 6**: Details on each outcome type, denoted by the variable `outcome_type` in the database. [a]This approach assumes a correlation coefficient of 0.5 between baseline and endpoint measurements. At this value, the standard deviations for change score and endpoint means are equal.

### Search Terms

#### Pubmed

**#1:** ("depress*"[Title/Abstract] OR "mood*"[Title/Abstract] OR "depression"[MeSH Terms] OR "mood disorders"[MeSH Terms] OR "depressive disorder"[MeSH Terms] OR "antidepress*"[Title/Abstract])
**#2:** ("psychedelic*"[Title/Abstract] OR "psilo*"[Title/Abstract] OR "Psilocybin"[MeSH Terms] OR "Psilocybe"[MeSH Terms] OR "magic mushroom*"[Title/Abstract] OR "teonanacatl"[Title/Abstract])
**#3:** ("Randomized Controlled Trial"[Publication Type] OR "Controlled Clinical Trial"[Publication Type] OR "Pragmatic Clinical Trial"[Publication Type] OR "Equivalence Trial"[Publication Type] OR "clinical trial, phase iii"[Publication Type] OR "Randomized Controlled Trials as Topic"[MeSH Terms] OR "Controlled Clinical Trials as Topic"[MeSH Terms] OR "Random Allocation"[MeSH Terms] OR "Double-Blind Method"[MeSH Terms] OR "Single-Blind Method"[MeSH Terms] OR "placebos"[MeSH Terms:noexp] OR "Control Groups"[MeSH Terms] OR

("random*"[Title/Abstract] OR "sham"[Title/Abstract] OR "placebo*"[Title/Abstract]) OR (("singl*"[Title/Abstract] OR "doubl*"[Title/Abstract]) AND ("blind*"[Title/Abstract] OR "dumm*"[Title/Abstract] OR "mask*"[Title/Abstract])) OR (("tripl*"[Title/Abstract] OR "trebl*"[Title/Abstract]) AND ("blind*"[Title/Abstract] OR "dumm*"[Title/Abstract] OR "mask*"[Title/Abstract])) OR ("control*"[Title/Abstract] AND ("study"[Title/Abstract] OR "studies"[Title/Abstract] OR "trial*"[Title/Abstract] OR "group*"[Title/Abstract])))
**#4: #1 AND #2 AND #3**

#### Embase

**#1:** ('depress*':ti,ab OR 'mood*':ti,ab OR 'depression'/exp OR 'mood disorders'/exp OR 'antidepress*':ti,ab)
**#2:** ('psychedelic*':ti,ab OR 'psilo*':ti,ab OR 'psilocybine'/exp OR 'psilocybe'/exp OR 'magic mushroom*':ti,ab OR 'teonanacatl':ti,ab)
**#3:** ('randomized controlled trial':it OR 'controlled clinical trial':it OR 'pragmatic clinical trial':it OR 'equivalence trial':it OR 'clinical trial, phase iii':it OR 'randomized controlled trial (topic)'/exp OR 'controlled clinical trial (topic)'/exp OR 'randomization'/exp OR 'double blind procedure'/exp OR 'single blind procedure'/exp OR 'placebo'/de OR 'control group'/exp OR ('random*':ti,ab OR 'sham':ti,ab OR 'placebo*':ti,ab) OR (('singl*':ti,ab OR 'doubl*':ti,ab) AND ('blind*':ti,ab OR 'dumm*':ti,ab OR 'mask*':ti,ab)) OR (('tripl*':ti,ab OR 'trebl*':ti,ab) AND ('blind*':ti,ab OR 'dumm*':ti,ab OR 'mask*':ti,ab)) OR ('control*':ti,ab AND ('study':ti,ab OR 'studies':ti,ab OR 'trial*':ti,ab OR 'group*':ti,ab)))
**#4: #1 AND #2 AND #3**

#### PsycInfo

**S1:** tiab("depress*" OR "mood*" OR "antidepress*") OR MAINSUBJECT.EXACT.EXPLODE("Depression (Emotion)") OR MAINSUBJECT.EXACT.EXPLODE("Major Depression") OR MAINSUBJECT.EXACT.EXPLODE("Affective Disorders")
**S2:** tiab("psychedelic*" OR "psilo*" OR "magic mushroom*" OR "teonanacatl") OR MAINSUBJECT.EXACT.EXPLODE("Psilocybin")
**S3:** ME("clinical trial") OR MAINSUBJECT.EXACT.EXPLODE("Randomized Clinical Trials") OR MAINSUBJECT.EXACT.EXPLODE("Randomized Controlled Trials") OR MAINSUBJECT.EXACT.EXPLODE("Clinical Trials") OR MAINSUBJECT.EXACT.EXPLODE("Random Sampling") OR MAINSUBJECT.EXACT.EXPLODE("Placebo") OR MAINSUBJECT.EXACT.EXPLODE("Experiment Controls") OR tiab("random*" OR "sham" OR "placebo*") OR tiab(("singl*" OR "doubl*" OR "tripl*" OR "trebl*") AND ("blind*" OR "dumm*" OR "mask*")) OR tiab("control*" AND ("study" OR "studies" OR "trial*" OR "group*"))
**#4: [S1] AND [S2] AND [S3]**

#### Web of Science

**#1:** (TI=depress* OR AB=depress*) OR (TI=mood* OR AB=mood*) OR TS=depression OR TS="mood disorders" OR TS="depressive disorder" OR (TI=antidepress* OR AB=antidepress*)
**#2:** (TI=psychedelic* OR AB=psychedelic*) OR (TI=psilo* OR AB=psilo*) OR TS=Psilocybin OR TS=Psilocybe OR (TI="magic mushroom*" OR AB="magic mushroom*") OR (TI=teonanacatl OR AB=teonanacatl)
**#3:** TS="Randomized Controlled Trial" OR TS="Controlled Clinical Trial" OR TS="Pragmatic Clinical Trial" OR TS="Equivalence Trial" OR TS="clinical trial, phase iii" OR TS="Randomized Controlled Trials as Topic" OR TS="Controlled Clinical Trials as Topic" OR TS="Random Allocation" OR TS="Double-Blind Method" OR TS="Single-Blind Method" OR TS=placebos OR TS="Control Groups" OR ((TI=random* OR AB=random*) OR (TI=sham OR AB=sham) OR (TI=placebo* OR AB=placebo*)) OR (((TI=singl* OR AB=singl*) OR (TI=doubl* OR AB=doubl*)) AND ((TI=blind* OR AB=blind*) OR (TI=dumm* OR AB=dumm*) OR (TI=mask* OR AB=mask*))) OR (((TI=tripl* OR AB=tripl*) OR (TI=trebl* OR AB=trebl*)) AND ((TI=blind* OR AB=blind*) OR (TI=dumm* OR AB=dumm*) OR (TI=mask* OR AB=mask*))) OR ((TI=control* OR AB=control*) AND ((TI=study OR AB=study) OR (TI=studies OR AB=studies) OR (TI=trial* OR AB=trial*) OR (TI=group* OR AB=group*)))
**#4: #1 AND #2 AND #3**

#### Scopus

**#1:** TITLE-ABS(depress*) OR TITLE-ABS(mood*) OR INDEXTERMS(depression) OR INDEXTERMS("mood disorders") OR INDEXTERMS("depressive disorder") OR TITLE-ABS(antidepress*)
**#2:** TITLE-ABS(psychedelic*) OR TITLE-ABS(psilo*) OR INDEXTERMS(Psilocybin) OR INDEXTERMS(Psilocybe) OR TITLE-ABS("magic mushroom*") OR TITLE-ABS(teonanacatl)
**#3:** INDEXTERMS("Randomized Controlled Trials as Topic") OR INDEXTERMS("Controlled Clinical Trials as Topic") OR INDEXTERMS("Random Allocation") OR INDEXTERMS("Double-Blind Method") OR INDEXTERMS("Single-Blind Method") OR INDEXTERMS(placebos) OR INDEXTERMS("Control Groups") OR (TITLE-ABS(random*) OR TITLE-ABS(sham) OR TITLE-ABS(placebo*)) OR ((TITLE-ABS(singl*) OR TITLE-ABS(doubl*)) AND (TITLE-ABS(blind*) OR TITLE-ABS(dumm*) OR TITLE-ABS(mask*))) OR ((TITLE-ABS(tripl*) OR TITLE-ABS(trebl*)) AND (TITLE-ABS(blind*) OR TITLE-ABS(dumm*) OR TITLE-ABS(mask*))) OR (TITLE-ABS(control*) AND (TITLE-ABS(study) OR TITLE-ABS(studies) OR TITLE-ABS(trial*) OR TITLE-ABS(group*)))
**#4: #1 AND #2 AND #3**

### Psychotherapy or Psychological Support

| Study | Psychotherapy Sessions and Frequency |
|---|---|
| Griffiths 2016 | 2 sessions between enrollment and baseline<br>1 session the day after each drug administration<br>2+ sessions between the first and second drug administration<br>2+ sessions between the second drug administration and the 6-month follow-up |
| Ross 2016 | 3 sessions before dose 1 (6 hours total)<br>3 sessions after dose 1 and before dose 2 (6 hours total)<br>3 sessions after dose 2 (6 hours total)<br>Continued ongoing support and integration with study therapist until 26 weeks |
| Davis 2021 | 6 to 8 hours of preparatory sessions with 2 facilitators between the baseline assessment and the day of dosing<br>A 7 to 10-hour dosing session conducted in a comfortable room under the supervision of the same facilitators<br>4 hours (2 sessions) of postdose integration sessions during which participants were invited to discuss their dosing experience with the facilitators (days 2 and 9) |
| Goodwin 2022 | At least 3 preparatory sessions before dosing<br>During administration sessions<br>2 integration sessions at day 2 and week 1 |
| Raison 2023 | 6 to 8 hours of preparatory sessions with 2 facilitators between the baseline assessment and the day of dosing<br>A 7 to 10-hour dosing session conducted in a comfortable room under the supervision of the same facilitators<br>4 hours (2 sessions) of postdose integration sessions during which participants were invited to discuss their dosing experience with the facilitators (days 2 and 9) |
| Von Rotz 2023 | 2 preparatory visits 4-6 days and 1 day before dosing (2 hours total)<br>1 session during drug administration (6 hours total)<br>3 integration visits 2, 8, and 14 days after dosing (3 hours total) |
| Back 2024 | 2 sessions before dosing at days -8 and -1<br>3 sessions after dosing at days 1, 8, and 15 |
| Rosenblat 2024 | 1 preparatory session before dosing; supportive therapy during dosing<br>2 integration sessions after dosing |
| Rieser 2025 | 2 sessions before dosing<br>3 sessions after dosing (1 day after dosing, 2 weeks after dosing, and 4 weeks after dosing, total of 5.5 hours) |
| Grob 2011 | unspecified |

| | |
|---|---|
| Carhart-Harris 2021 | 1 session one day before first dose<br>1 session one day after each dose<br>1 session during each dose<br>3 phone sessions in the three week period after each dose<br>Optional aftercare support following the 6 week primary outcome |
| Krempien 2023 | unspecified |

**eTable 7**: Psychotherapy or psychological support sessions and their frequency for each study.

### Excluded Reports

| **Excluded Study Title:** | **Excluded Study Author and Year:** | **Reason for Exclusion:** |
|---|---|---|
| Preliminary safety and effectiveness of psilocybin-assisted therapy in adults with fibromyalgia: an open-label pilot clinical trial | Aday 2025[7] | Not an RCT |
| In patients with major depressive disorder, psilocybin administration is associated with reduced amygdala response to negative affective stimuli and normalization of cortical glutamate one week after psilocybin, and improved cognitive flexibility one and four weeks after psilocybin | Barrett 2019[8] | Re-analysis/Secondary report |
| Subjective effects of psilocybin predict next-day differences in default mode network and medial temporal lobe functional connectivity | Barrett 2016[9] | Wrong patient population (e.g., healthy, pediatric) |
| Effects of Psilocybin on Suicidal Ideation in Patients with Life-Threatening Cancer | Benville 2021[10] | Re-analysis/Secondary report |
| Mood, craving, and self-efficacy in psilocybin-assisted treatment of alcoholism | Bogenschutz 2015[11] | Wrong outcomes/no mood outcomes |
| Clinical Interpretations of Patient Experience in a Trial of Psilocybin-Assisted Psychotherapy for Alcohol Use Disorder | Bogenschutz 2018[12] | Wrong outcomes/no mood outcomes |
| Percentage of Heavy Drinking Days Following Psilocybin-Assisted Psychotherapy vs. Placebo in the Treatment of Adult Patients with Alcohol Use Disorder: A Randomized Clinical Trial | Bogenschutz 2022[13] | Wrong outcomes/no mood outcomes |

| | | |
|---|---|---|
| Do magic mushrooms help against depression? Controlled intake of psilocybin in pilot study is effective | Bruhn 2016 | Record not retrieved |
| Comparing Antidepressant Effects of Psilocybin-Assisted Psychotherapy in Individuals That Were Unmedicated at Initial Screening Versus Individuals Discontinuing Medications for Study Participation: Comparaison des effets antidépresseurs de la psychothérap | Chisamore 2025[14] | Re-analysis/Secondary report |
| Predicting the outcome of psilocybin treatment for depression from baseline fMRI functional connectivity | Copa 2024[15] | Re-analysis/Secondary report |
| Acute Psychedelic Effects and Clinical Outcomes are Associated with Therapeutic Alliance Among Participants Receiving Psilocybin Therapy | Davis 2024[16] | Re-analysis/Secondary report |
| Increased global integration in the brain after psilocybin therapy for depression | Daws 2022[17] | Re-analysis/Secondary report |
| Different hierarchical reconfigurations in the brain by psilocybin and escitalopram for depression | Deco 2024[18] | Re-analysis/Secondary report |
| Psilocybin therapy increases cognitive and neural flexibility in patients with major depressive disorder | Doss 2021[19] | Re-analysis/Secondary report |
| Psilocybin therapy for treatment resistant depression: prediction of clinical outcome by natural language processing | Dougherty 2023[20] | Re-analysis/Secondary report |
| Treatment Expectancies and Psilocybin vs. Escitalopram for Depression | Dutcher 2025[21] | Re-analysis/Secondary report |
| Music as a collaborating actor: new insights into the nature and role of music in psychedelic-assisted psychotherapy | Dwyer 2025[a22] | Re-analysis/Secondary report |
| Effect of psilocybin versus escitalopram on depression symptom severity in patients with moderate-to-severe major depressive disorder: observational 6-month follow-up of a phase 2, double-blind, randomised, controlled trial | Erritzoe 2024[23] | Re-analysis/Secondary report |
| Dose-Dependent Acute Subjective | Goodwin 2022[24] | Re-analysis/Secondary |

| | | |
|---|---|---|
| Psychedelic Effects Following COMP360 Psilocybin Across Three Clinical Studies and its Relationship to Therapeutic Response | | report |
| The role of the psychedelic experience in psilocybin treatment for treatment-resistant depression | Goodwin 2025[25] | Re-analysis/Secondary report |
| Predicting Depression Outcomes Through the Influence of Therapeutic Alliance and the Psychedelic Experience Using Path Modelling in a Phase IIb Randomized Controlled Trial of COMP360 Psilocybin Therapy | Goodwin 2022[26] | Re-analysis/Secondary report |
| A single dose of psilocybin produces substantial and enduring decreases in anxiety and depression in patients with a life-threatening cancer diagnosis: A randomized double-blind trial | Griffiths 2015[27] | Conference abstract - full text already included |
| Psilocybin-assisted treatment of major depressive disorder: Results from a randomized trial | Griffiths 2019[28] | Conference abstract - full text already included |
| Psilocybin treatment for anxiety in patients with advanced-stage cancer | Grob 2012[29] | Conference abstract - full text already included |
| Psilocybin-Assisted Therapy for MDD: Current Evidence and Clinical Considerations | Gukasyan 2022[30] | Conference abstract - full text already included |
| Dissociable effects of psilocybin and escitalopram for depression on processing of musical surprises | Harding 2025[31] | Re-analysis/Secondary report |
| Nature-themed video intervention may improve cardiovascular safety of psilocybin-assisted therapy for alcohol use disorder | Heinzerling 2023[32] | Not an RCT |
| The effects of psilocybin therapy versus escitalopram on cognitive bias: A secondary analysis of a randomized controlled trial. | Henry 2025[33] | Re-analysis/Secondary report |
| Single-dose psilocybin therapy for alcohol use disorder: Pharmacokinetics, feasibility, safety and efficacy in an open-label study | Jensen 2025[34] | Not an RCT |
| Assessing Cognitive Outcomes in | Johnson 2025[35] | Re-analysis/Secondary |

| | | |
|---|---|---|
| Treatment-Resistant Depression Following Psilocybin-Assisted Psychotherapy | | report |
| Psilocybin increases emotional empathy in patients with major depression | Jungwirth 2025[36] | Re-analysis/Secondary report |
| Changes in Symptoms of Anhedonia With Psilocybin-Assisted Psychotherapy: A Secondary Analysis of a Randomized Clinical Trial for Treatment-Resistant Depression | Kaczmarek 2025[37] | Re-analysis/Secondary report |
| A machine learning using NLP to predict long-term patient response | Kumar 2025[38] | Re-analysis/Secondary report |
| The therapeutic alliance between study participants and intervention facilitators is associated with acute effects and clinical outcomes in a psilocybin-assisted therapy trial for major depressive disorder. | Levin 2024[39] | Re-analysis/Secondary report |
| Psilocybin-Assisted Group Psychotherapy + Mindfulness Based Stress Reduction (MBSR) for Frontline Healthcare Provider COVID-19 Related Depression and Burnout: A Randomized Clinical Trial. | Lewis 2025[40] | Pre-print/Conference poster |
| Improving Workspace and Psychosocial Function in Treatment-Resistant Depression: Exploratory Outcomes From an Open-Label Feasibility Trial of Psilocybin-Assisted Psychotherapy | Lipsitz 2025[41] | Re-analysis/Secondary report |
| The impact of antidepressant discontinuation prior to treatment with psilocybin for treatment-resistant depression. | Marwood 2024[42] | Re-analysis/Secondary report |
| Efficacy and safety of psilocybin in treatment-resistant major depression (EPIsoDE) - study design, rationale and current status | Mertens 2021[43] | Protocol/incomplete study |
| Psilocybin for treatment-resistant major depression - Results from an academic phase 2B randomized-controlled trial | Mertens 2024[44] | Pre-print/Conference poster |
| Therapeutic mechanisms of psychedelic drugs: Changes in amygdala and prefrontal functional connectivity during emotional processing after psilocybin for | Mertens 2019[45] | Not an RCT |

| | | |
|---|---|---|
| treatment-resistant depression | | |
| Methodological challenges in psychedelic drug trials: Efficacy and safety of psilocybin in treatment-resistant major depression (EPISODE) - rationale, study design and current status | Mertens 2021[46] | Protocol/incomplete study |
| Therapeutic Alliance and Rapport Modulate Responses to Psilocybin Assisted Therapy for Depression | Murphy 2022[47] | Re-analysis/Secondary report |
| A Bayesian Reanalysis of a Trial of Psilocybin versus Escitalopram for Depression | Nayak 2023[48] | Re-analysis/Secondary report |
| Psilocybin works well against depression | Ostensjo 2024 | Record not retrieved |
| Psilocybin-induced changes in neural reactivity to alcohol and emotional cues in patients with alcohol use disorder: an fMRI pilot study | Pagni 2024[49] | Wrong outcomes/no mood outcomes |
| Multidimensional Personality Changes Following Psilocybin-Assisted Therapy in Patients with Alcohol Use Disorder: Results from a Double-Blind, Placebo-Controlled Clinical Trial | Pagni 2025[50] | Wrong outcomes/no mood outcomes |
| Spontaneous changes in cigarette smoking following psilocybin-assisted treatment for alcohol use disorder | Pagni 2024[51] | Wrong outcomes/no mood outcomes |
| The effects of psilocybin on the dynamics of EEG changes in human volunteers | Pálenícek 2018[52] | Wrong patient population (e.g., healthy, pediatric) |
| Psilocybin-assisted psychotherapy improves psychiatric symptoms across multiple dimensions in patients with cancer | Petridis 2024[53] | Re-analysis/Secondary report |
| Psilocybin Assisted Therapy for Treatment-Resistant Depression: A Phase II, Randomized, Feasibility Study | Rosenblat 2022[54] | Conference abstract - full text already included |
| Psilocybin-assisted psychotherapy for treatment-resistant depression: A phase II, randomized, feasibility study | Rosenblat 2023[55] | Conference abstract - full text already included |
| Acute and Sustained Reductions in Loss of Meaning and Suicidal Ideation Following | Ross 2021[56] | Re-analysis/Secondary report |

| | | |
|---|---|---|
| Psilocybin-Assisted Psychotherapy for Psychiatric and Existential Distress in Life-Threatening Cancer | | |
| Exploratory Controlled Study of the Migraine-Suppressing Effects of Psilocybin | Schindler 2021[57] | Wrong outcomes/no mood outcomes |
| Klinische erfahrungen mit psilocybin (CY 39 sandoz) | Sercl 1961[58] | non-English language |
| Changes in music-evoked emotion and ventral striatal functional connectivity after psilocybin therapy for depression | Shukuroglou 2023[59] | Not an RCT |
| Sub-acute effects of psilocybin on EEG correlates of neural plasticity in major depression: Relationship to symptoms. | Skosnik 2023[60] | Not an RCT |
| Psilocybin-assisted therapy for major depressive disorder: An exploratory placebo-controlled, fixed-order trial. | Sloshower 2023[61] | Not an RCT |
| Psychological flexibility as a mechanism of change in psilocybin-assisted therapy for major depression: results from an exploratory placebo-controlled trial | Sloshower 2024[62] | Not an RCT |
| Psilocybin to promote synaptogenesis in the brains of patients with mild cognitive impairment | Song 2023[63] | Protocol/incomplete study |
| Psilocybin assisted psychotherapy for patients with ovarian cancer: A subset analysis of a randomized, double-blind, placebo-controlled, phase II clinical trial | Spinosa 2024[64] | Re-analysis/Secondary report |
| Enhanced visual contrast suppression during peak psilocybin effects: Psychophysical results from a pilot randomized controlled trial. | Swanson 2024[65] | Wrong patient population (e.g., healthy, pediatric) |
| Assessing expectancy and suggestibility in a trial of escitalopram v. psilocybin for depression. | Szigeti 2024[66] | Re-analysis/Secondary report |
| Reduced Brain Responsiveness to Emotional Stimuli With Escitalopram But Not Psilocybin Therapy for Depression. | Wall 2025[67] | Re-analysis/Secondary report |
| Reduced brain responsiveness to emotional stimuli with escitalopram but not psilocybin | Wall 2023[68] | Re-analysis/Secondary report |

| | | |
|---|---|---|
| therapy for depression | | |
| A critical evaluation of QIDS-SR-16 using data from a trial of psilocybin therapy versus escitalopram treatment for depression | Weiss 2023[69] | Re-analysis/Secondary report |
| Unique psychological mechanisms underlying Psilocybin Therapy versus Escitalopram Treatment in the treatment of major depressive disorder | Weiss 2024[70] | Re-analysis/Secondary report |
| How does psilocybin therapy work? An exploration of experiential avoidance as a putative mechanism of change. | Zeifman 2023[71] | Re-analysis/Secondary report |
| Dynamic Mediodorsal Thalamus Activity Predicts and Reflects Longitudinal Treatment Outcomes in Psilocybin Therapy for Treatment-Resistant Depression | Zhang 2025[72] | Not an RCT |
| Changes in Resting State Functional Connectivity After Psilocybin for Body Dysmorphic Disorder | Zhu 2023[73] | Not an RCT |
| Single-dose psilocybin alters resting state functional networks in patients with body dysmorphic disorder | Zhu 2025[74] | Not an RCT |

**eTable 8**: List of reports excluded during full-text review. Excluded secondary reports also appear in the next table. [a]The authors of Dwyer 2025 were contacted regarding depression outcomes for this study, however the publication was not yet available.

### Merged and Excluded Secondary Reports

| **Primary study:** | **Merged report:** | **What does the merged report(s) add to the database?** | **Additional secondary reports that were excluded in full-text review:** |
|---|---|---|---|
| Griffiths 2016[75] | N/A | N/A | N/A |
| Ross 2016[76] | Agin-Liebes 2020[a77] | N/A | Ross 2021[56]<br>Benville 2021[10]<br>Spinosa 2024[64] |
| Davis 2021[78] | Gukasyan 2022[a79]<br>Correction to Davis 2021[80] | N/A | Barrett 2019[8]<br>Doss 2021[19]<br>Levin 2024[39] |

| | | | |
|---|---|---|---|
| | | | Davis 2024[16] |
| Goodwin 2022[81] | Goodwin 2023[82]<br>Goodwin 2023[a83]<br>Goodwin 2025[a84] | QIDS-SR-16 | Goodwin 2022a[26]<br>Goodwin 2022b[24]<br>Dougherty 2023[20]<br>Marwood 2024[42]<br>Goodwin 2025[25]<br>Kumar 2025[85] |
| von Rotz 2023[86] | Correction to von Rotz 2023[87] | N/A | Jungwirth 2025[36] |
| Raison 2023[88] | Correction to Raison 2023[89] | N/A | N/A |
| Rosenblat 2024[90] | N/A | N/A | Chisamore 2025[14]<br>Johnson 2025[35]<br>Kaczmarek 2025[37]<br>Lipsitz 2025[41] |
| Back 2024[91] | Correction to Back 2024[92] | N/A | N/A |
| Rieser 2025[93] | N/A | N/A | N/A |
| Grob 2011[94] | N/A | N/A | N/A |
| Carhart-Harris 2021[95] | Barba 2022[a96]<br>Weiss 2024[a97]<br>Correction to Weiss 2024[98] | N/A | Daws 2022[17]<br>Murphy 2022[47]<br>Nayak 2023[48]<br>Weiss 2023[69]<br>Wall 2023[68]<br>Zeifman 2023[71]<br>Copa 2024[15]<br>Deco 2024[18]<br>Erritzoe 2024[23]<br>Szigeti 2024[66]<br>Weiss 2024[70]<br>Dutcher 2025[21]<br>Harding 2025[31]<br>Henry 2025[33]<br>Wall 2025[67] |
| Krempien 2023[99] | CYBIN January 2025 Corporate Slide Deck (sypres.io/assets/pdfs/CYBN-Corporate-Deck-January-2025-Website-Final.pdf) | MADRS (supplemented via author contact) | N/A |

**eTable 9**: Record of secondary reports that were either merged with the parent study for extraction or excluded in full-text screening. [a]Information extracted from these reports either did

not relate to depression or was not in a format conducive to between-arm meta-analysis and therefore did not make it into the publicly released database.

### Record of Contacting Study Authors

| Study | What info was requested from authors? | What information was provided from authors? |
|---|---|---|
| Ross 2016 | Data mean, SE and N for data reported only in figures:<br>● For the HADS Total, HADS-A, HADS-D, BDI, and STAI: Baseline, 1 day pre-dose 1, 1 day post-dose 1, 2 weeks post-dose 1, 6 weeks post-dose 1, 7 weeks post-dose 1 and 1 day pre-dose 2, 1 day post-dose 2, 6 weeks post-dose 2 for both psilocybin-first and niacin-first arms<br>● For response and remission outcomes: 2-4 weeks pre-dose 1, 1 day post-dose 1, 7 weeks post-dose 1 and 1 day pre-dose 2, 26 weeks post-dose 2, long-term follow-up data (4.5 years after psilocybin dose) | N/A (no response received) |
| Griffiths 2016 | Clarification on N at the high-dose 1st post-session 1 for different outcome measures (caption on table in paper says ‘25 or 26’):<br>● GRID-HAMD-17<br>● BDI<br>● HADS Depression<br>● HAM-A<br>● STAI-Trait Anxiety<br>● POMS | Clarification on N for outcome measures:<br>● N=25 participants for GRID-HAMD and HAM-A<br>● N=18 participants for LAP-R Death Transcendence (they started administering it part way through the study; verified that |

| | | |
|---|---|---|
| | • BSI<br>• MQOL<br>• LAP-R<br>• LOT-R | mean (SEM) reported in Table 4 is accurate for this sample size<br>• N=26 participants for all other outcomes (BDI, HADS-D, STAI State, STAI Trait, POMS, BSI, MQOL, LAP-R, LOT-R) |
| | Question about assessor blinding/independence: Who performed the GRID-HAMD-17 clinical assessment?<br>• third-party independent raters<br>• independent study staff<br>• non-independent study staff (involved in the administration of therapy or other aspects of the trial involving patient contact)<br>• other (explain)<br>Note: Also please indicate whether raters were blinded or unblinded to participant condition (intervention vs control) or you have any additional elaboration. | Non-independent study staff, who were blinded to the dosing condition of each participant. |
| Davis 2021 | Who performed clinical assessments?<br>• third-party independent raters<br>• independent study staff<br>• non-independent study staff (involved in the administration of therapy or other aspects of the trial involving patient contact)<br>• other (explain)<br>Note: Also please indicate whether raters were blinded or unblinded to participant condition (intervention vs control) or you have any additional elaboration. | The raters were affiliated with JHU but not otherwise involved in any aspect of the trial. e.g. Independent study staff. |

| | | |
|---|---|---|
| | Was the analysis ITT or PP? | PP |
| Goodwin 2022 | Unadjusted post-intervention mean, SD, and N for the following timepoints and outcomes:<br>● MADRS: Day 2, Week 1, Week 3, Week 6, Week 9, and Week 12 for the 25 mg, 10 mg, and 2 mg arms<br>● QIDS-SR: Week 3 and Week 12 for the 25 mg, 10 mg, and 1 mg arms<br>● SDS: Week 3 and Week 12 for the 25 mg, 10 mg, and 1 mg arms<br>● WSAS: Week 3 and Week 12 for the 25 mg, 10 mg, and 1 mg arms<br>● EQ-5D-3L: Week 3 and Week 12 for the 25 mg, 10 mg, and 1 mg arms<br>● GAD-7: Week 3 and Week 12 for the 25 mg, 10 mg, and 1 mg arms<br>● EQ-VAS: Week 3 and Week 12 for the 25 mg, 10 mg, and 1 mg arms<br>● PANAS Positive: Day 2 and Week 3 for the 25 mg, 10 mg, and 1 mg arms<br>● PANAS Negative: Day 2 and Week 3 for the 25 mg, 10 mg, and 1 mg arms<br>● DSST: Day 2, Week 3, and Week 12 for the 25 mg, 10 mg, and 1 mg arms | The authors reported they are unable to share any additional data prior to NDA submission. |

| | | |
|---|---|---|
| von Rotz 2023 | Question about assessor blinding/independence: Who performed the MADRS clinical assessment?<br>• third-party independent raters<br>• independent study staff<br>• non-independent study staff (involved in the administration of therapy or other aspects of the trial involving patient contact)<br>• other (explain)<br>Note: Also please indicate whether raters were blinded or unblinded to participant condition (intervention vs control) or you have any additional elaboration. | Non-independent study staff involved in the administration of the therapy. |
| Raison 2023 | Unadjusted post-intervention mean, SD, and N for the following timepoints and outcomes, with particular interest in post-intervention values for the MADRS at Day 43:<br>● MADRS: Day 2, Day 8, Day 15, Day 29, and Day 43 for both psilocybin and niacin arms<br>● SDS: Day 8, Day 15, Day 29, and Day 43 for both psilocybin and niacin arms<br>● HAM-A: Day 8, Day 15, Day 29, and Day 43 for both psilocybin and niacin arms<br>● Q-LES-Q: Day 8, Day 15, Day 29, and Day 43 for both psilocybin and niacin arms<br>● ODQ: Day 43 for both psilocybin and niacin arms<br>● SMDDS: Day 8 and Day 43 for both psilocybin and niacin | N/A (no response received) |

| | | |
|---|---|---|
| | arms | |
| Rosenblat 2024 | Unadjusted post-intervention mean, SD, and N for the following timepoints and outcomes, with particular interest in post-intervention values for the MADRS at Day 14, and the QIDS-SR at Day 14:<br>● **MADRS**: Day -14, Day 0, Day 1, Day 7, and Day 14 for both immediate and delayed treatment groups<br>● **QIDS-SR:** Day -14, Day 0, Day 1, Day 7, and Day 14 for both immediate and delayed treatment groups<br>● CGI: Day -14, Day 0, Day 1, Day 7, and Day 14 for both immediate and delayed treatment groups<br>● MADRS Suicidal Ideation Item 10: Day -14, Day 0, Day 1, Day 7, and Day 14 for both immediate and delayed treatment groups<br>● GAD-7: Day 0 and Day 14 for both immediate and delayed treatment groups | Day 14 mean, SD, and N for the MADRS. |
| | Clarification on relationship of the clinician raters (those who performed the MADRS assessment) with the trial participants: (i.e., were they: third-party independent raters; independent study staff; non-independent study staff involved in the | Non-independent study staff who weren't involved in the administration of therapy but were involved in other aspects of the trial involving patient contact. |

| | | |
|---|---|---|
| | administration of therapy or other aspects of the trial involving patient contact; or other) | |
| Back 2024 | N/A | N/A |
| Rieser 2025 | Raw, unadjusted post-intervention mean, SD, and N for the BDI for the following timepoints: Day -14, Day -6, Day 1, Day 14, Day 30, Day 96, and Day 190 for both placebo and psilocybin groups<br><br>Why was mannitol selected as the comparator - is it a standard placebo, or does it produce any physiological effects like flushing or increased heart rate in the same way that niacin placebos produce?<br><br>Did you collect any clinician-administered scales not included in the paper? | Raw, unadjusted post-intervention mean, SD, and N for the BDI for the following timepoints: Day -14, Day -6, Day 1, Day 14, Day 30, Day 96, and Day 190 for both placebo and psilocybin groups<br><br>Mannitol has no known psychoactive effects or impact on heart rate and has been used in previous studies by this group. They did not observe any reports of increased urination.<br><br>There were no clinician-administered mood scales collected. |
| Grob 2011 | Numerical data provided in figures:<br>● BDI: 1 day before the experimental session, 1 day after the experimental session, 2 weeks after the experimental session, 6-month follow-up for both data separated by intervention and control groups (all data in Figure 3a) and data collapsed across groups (all data in Figure 3b)<br>● POMS and STAI: 1 day before the experimental session, | N/A (no response received) |

| | | |
|---|---|---|
| | <ul><li>6 hours after the dose/at the conclusion of the experimental session, 6-month follow-up for both data separated by intervention and control groups (all data in Figures 4a and 5a), and data collapsed across all groups (all data in Figure 4b and 5b)</li></ul><br>Raw data from Brief Psychiatric Rating Scale outcome measure<br><br>Any further details about the baseline characteristics of the population, detailed inclusion/exclusion criteria, PRISMA diagram or information detailing the withdrawals<br><br>Clarification as to whether figure 3a provided pre-crossover or post-crossover data.<br><br>Request for pre-crossover data (mean, SE, and N):<ul><li>BDI: 1 day before first experimental session and 2 weeks after the first experimental session, for both the psilocybin-first group and niacin-first group separately</li></ul> | N/A (no response received)<br><br>N/A (no response received)<br><br>N/A (no response received)<br><br>N/A (no response received) |
| Carhart-Harris 2021 | Unadjusted continuous outcomes from Table 2 in NEJM as N, mean, and SD rather than change scores. If it is overly burdensome to share all of the outcomes, please prioritize the depression-related and STAI. | Unadjusted endpoint values for:<ul><li>QIDS-SR-16</li><li>BDI-1A</li><li>HAM-D-17</li><li>MADRS</li><li>STAI</li></ul> |

| | | |
|---|---|---|
| | Could you please let us know which of the categories below the assessment raters belong in?<br>● third-party independent raters<br>● independent study staff<br>● non-independent study staff (involved in the administration of therapy or other aspects of the trial involving patient contact)<br>● other (explain)<br>Also please indicate whether raters were blinded or unblinded to participant condition (psilo vs ssri) or you have any additional elaboration. | Non-independent study staff who were not involved in therapy but who were involved in other aspects of the trial involving patient contact such as screening for some participants. |
| Krempien 2023 | Clarification on different reports of the N for each arm<br><br>Unadjusted raw values for MADRS for each arm (12 mg, 16 mg, and placebo) at the following time points: Baseline, Day 21<br><br>What is the “drug” for the placebo condition?<br><br>Was any self-report depression data collected (such as BDI or QIDS)?<br><br>Who performed the clinical assessments in the study? (i.e., third-party independent raters, independent study staff, non-independent study staff involved in the administration of therapy or other aspects of the trial | Clarification on different reports of the N for each arm<br><br>Unadjusted raw values for MADRS for each arm (12 mg, 16 mg, and placebo) at the following time points: Baseline, Day 21<br><br>N/A (no response received)<br><br>N/A (no response received)<br><br>N/A (no response received) |

| | |
|---|---|
| | involving patient contact, or other). Were raters blinded or unblinded to participant condition (intervention vs. control)? |

**eTable 10**: Record of author contact and responses via email.

### Risk of Bias Assessment Standard Operating Procedure

Standard Operating Procedure for using Cochrane's RoB2 tool. See the *Risk of Bias* section in the supplementary methods for more general details and discussion.

#### Overall Workflow

| **Notes:** |
|---|
| 1. Each reviewer completes the RoB2 tool Excel spreadsheet, using the descriptions in the tool as well as this SOP for guidance.<br>2. Discuss and create a consensus version of the RoB2 tool Excel spreadsheet.<br>3. Create a new spreadsheet from the consensus spreadsheet that includes only overall D1-D5 results for use in R packages to create tables/graphs for RoB.<br>*Always fill out the ITT version of the form, even if the study uses PP analysis.<br><br>*If we override the algorithm in the spreadsheet for any reason, our workflow is to: 1) discuss; 2) document in Notion and the Excel spreadsheet |

#### Domain 1 - Randomization

+ **1.1** - Was the allocation sequence random?
+ **1.2** - Was the allocation sequence concealed until participants were enrolled and assigned to interventions?
+ **1.3** - Did baseline differences between intervention groups suggest a problem with the randomization process?
    + Integrate metapsy tool: If no p-values are provided in the baseline characteristics table for the study you are examining, or if you are uncertain whether the differences are statistically significant, input means, standard deviations, etc. into this tool to determine if baseline differences between intervention groups are significant.

+ Most of the time, the answer to this question will be "no," unless there are substantial differences in the baseline outcome measure between groups. You only need to check the outcome measure (i.e., MADRS, BIDS) - there is no reason to think that age or sex imbalance, for example, would severely influence the results.

#### Domain 2 - Deviations

+ *The key assumption we are making for this domain is that it is meant to address major protocol deviations more specifically, not dropout/missingness, which is the concern of Domain 3.*
+ **2.1** - Were participants aware of their assigned intervention during the trial?
    + The answer will probably be "PY" (due to side effects of psychedelics) unless the study actually assesses blinding of participants, and from their assessment you can confidently mark 'Y' if participants were definitely aware of their assigned intervention or 'N' if they were not.
+ **2.2** - Were carers and people delivering the interventions aware of participants' assigned intervention during the trial?
    + The answer will probably be "PY" (due to side effects of psychedelics) unless the study actually assesses blinding of staff and you can confidently mark 'Y' or 'N'.
+ **2.3** - Were there deviations from the intended intervention that arose because of the trial context?
    + Try not to put 'NI' for this even if information is sparse - it actually makes a difference in the algorithm.
    + Things outside of trial context (i.e., participant dropout or restarting antidepressants) usually do not count here. Only count starting antidepressants again if there is a solid reason that one group had a higher rate of starting up antidepressants.
    + An example of what counts as a deviation due to trial context is if one arm received greater levels of psychotherapy compared to the other arm due to safety concerns.
+ **2.4** - Were these deviations likely to have affected the outcome?
    + You can answer 'N' if ITT and PP analyses were consistent.
+ **2.5** - Were these deviations from intended intervention balanced between groups?
+ **2.6** - Was an appropriate analysis used to estimate the effect of assignment to intervention?
    + If it is unspecified whether ITT or PP analysis was used, you can put 'PN'.
+ **2.7** - Was there potential for a substantial impact (on the result) of failure to analyze participants in the group to which they were randomized?
    + Cochrane states for this question that "it is not possible to specify a precise rule." Usually, this question will be PN/N, unless there is evidence of substantial impact.
+ For studies that use PP analysis, but have low/balanced deviations or if the deviations described occur after the primary endpoint that is being assessed, the overall risk of bias

for this domain can be overridden to ‘Low’ rather than the algorithm’s assessment of ‘Some concerns’.

#### Domain 3 - Missing Data/Dropout

+ **3.1** - Were data for this outcome available for all, or nearly all, participants randomized?
    + Imputed data still counts as missing data.
    + Calculate: for continuous outcomes, 95% counts as 'nearly all'.
+ **3.2** - Is there evidence that the result was not biased by missing outcome data?
    + If dropouts are low and/or balanced between the two groups, the answer will probably be PY/Y.
    + If dropouts are high and/or unequal between the two groups (especially if related to adverse events/severity of symptoms), and robust sensitivity analyses were not performed to test if there is something different about the groups at baseline or other timepoints, then the answer will probably be PN/N.
+ **3.3** - Could missingness in the outcome depend on its true value?
    + This will be 'PY' if the main reason people dropped out was due to responding poorly (or really well) to treatment.
    + Most of the time, the answer to this question will be 'Y/PY' unless the documented reasons for dropout were all unrelated to health status/outcome.
+ **3.4** - Is it likely that missingness in the outcome depended on its true value?
    + Usually, the answer to this question will be 'PN'. Only put 'Y' for this question unless it is well-documented that missingness in the outcome depended on its true value (i.e., if all the dropouts were due to increased suicidality, or if all participants in the psilocybin group dropped out because their depression was resolved and they no longer felt the need to continue the trial).

#### Domain 4 - Measurements

+ **4.1** - Was the method of measuring the outcome inappropriate?
    + Usually 'N'.
+ **4.2** - Could measurement or ascertainment of the outcome have differed between intervention groups?
    + Usually 'N'.
+ **4.3** - Were outcome assessors aware of the intervention received by study participants?
    + In some cases, it might be necessary to contact study authors, i.e., if the outcome is clinician-administered (i.e., MADRS), and the study does not specify who measured the outcome.
    + Clinician-administered outcomes where blinded third-party or independent study staff raters performed the measurement receive a ‘N’.
    + If the authors do not respond, it is fair to put 'NI' here and 'PY' for 4.4.
    + Self-report outcome measurements get a 'PY' here.
+ **4.4** - Could assessment of the outcome have been influenced by knowledge of intervention received?

    + For self-report, if there is no evidence that participants under-reported, this will be 'PY.'
    + For clinician-administered outcomes with unblinded/functionally unblinded study staff, this will be 'PY'.
+ **4.5** - Is it likely that the assessment of the outcome was influenced by knowledge of intervention received?
    + For self-report, this question will be 'PN' unless there is substantial evidence or reason to believe that participants underreported their symptoms.
    + For clinician-administered outcomes by non-independent study staff, a judgement call must be made based on their level of patient contact, potential for functional unblinding, and overall involvement in the study.
    + For clinician-administered outcomes in an open-label study assessed by explicitly unblinded non-independent study staff, this will be a 'PY'.

#### Domain 5 - Pre-registration

+ **5.1** - Were the data that produced these results analyzed in accordance with a pre-specified analysis plan that was finalized before unblinded outcome data were available for analysis?
    + Look at the trial protocol/statistical analysis plan if provided.
    + Look at clinicalTrials.gov (making sure to examine the version history).
+ **Is the numerical result being assessed likely to have been selected, on the basis of the results, from...**
+ **5.2** - ...multiple eligible outcome measurements (e.g., scales, definitions, timepoints) within the outcome domain?
+ **5.3** - ...multiple eligible analyses of the data?
